# Computational phenotyping of aberrant belief updating in individuals with schizotypal traits and schizophrenia

**DOI:** 10.1101/2023.11.27.23299069

**Authors:** Nace Mikus, Claus Lamm, Christoph Mathys

## Abstract

Psychotic and psychotic-like experiences are thought to emerge from various patterns of disrupted belief updating. These include belief rigidity, overestimating the reliability of sensory information, and misjudging task volatility. Yet, these substrates have never been jointly addressed under one computational framework and it is not clear to what degree they reflect trait-like computational patterns. Here, we introduce a computational model that describes interindividual differences in how individuals update their beliefs about a volatile environment through noisy observations and use this model in a series of studies. In a test-retest study with healthy participants (N = 45, 4 sessions), we find that interclass correlations were moderate to high for session-level model parameters and excellent for averaged belief trajectories and learning rates estimated through hierarchical Bayesian inference. Across three studies (total N=590) we then demonstrate that two distinct computational patterns describe two different transdiagnostic categories. Higher uncertainty about the task volatility is related to schizotypal traits (N = 45, d = 0.687, P = 0.02, N = 437, d = 0.14, P = 0.032) and to positive symptoms in a sample of patients with schizophrenia (N = 108, d = 0.187, P = 0.039), when learning to gain rewards. In contrast, depressive-anxious traits were associated with more rigid beliefs about the underlying mean (N = 437, d = −0.125, P = 0.006) and outcome variance (N = 437, d = −0.111, P = 0.013), as were negative symptoms in patients with schizophrenia (d = - 0.223, P = 0.026, d = −0.298, P = 0.003), when learning to avoid losses. These findings suggest that individuals high on schizotypal traits across the psychosis continuum are less likely to learn or utilize higher-order statistical regularities of the environment and showcase the potential of clinically relevant computational phenotypes for differentiating symptom groups in a transdiagnostic manner.

## Introduction

A computational phenotype is a computational parameter that provides a trait-like, mechanistic description of a psychological, behavioral, or neural process. As such, computational phenotypes can enable transdiagnostic assessments of psychiatric symptoms that span many levels of analysis and have the potential to guide psychiatric treatment (Paulus, Huys, & Maia, 2016; Stephan & Mathys, 2014). One area where computational approaches have proven particularly promising is in describing neurocomputational mechanisms of how humans and non-human animals infer latent unobservable states from sensory observations. In particular, a rich body of theoretical and empirical work supports the notion that schizotypal traits across the severity spectrum can be cast as aberrant inference or learning (Adams, Stephan, Brown, Frith, & Friston, 2013; Corlett, Frith, & Fletcher, 2009; Fletcher & Frith, 2009; Stephan, Baldeweg, & Friston, 2006).

Under normative accounts of statistical inference, the degree to which new observations reflect meaningful occurrences in our environment should depend on several distinct sources of uncertainty, such as how reliable we feel our observation was (outcome uncertainty), or how likely we believe such an occurrence to be (environmental uncertainty). This weighting of relative uncertainties (or their inverses, precisions) when integrating sensory information with prior expectations has been referred to as precision-weighted belief updating (Friston, 2009; Mathys, Daunizeau, Friston, & Stephan, 2011). Overestimating either the precision of sensory sensations or the probability of unlikely events can both lead to attributing too much importance to random chance occurrences. Within this framework, the computational substrate underlying psychotic symptoms has been suggested to arise from this aberrant attribution of salience to internal sensations and random external events, leading to a psychological state where the world seems imbued with significant experiences that provoke bizarre explanations (Fletcher & Frith, 2009; Jaspers, 1997; Kapur, 2003).

The challenge of separating outcomes arising from noise in observation from outcomes that reflect latent environmental changes has been operationalized in probabilistic reversal learning tasks. Empirical research using various variations of learning tasks has shown that patients with schizophrenia tend to update their beliefs more rapidly (Garety, Hemsley, & Wessely, 1991; Morawetz, Bode, Baudewig, Jacobs, & Heekeren, 2016), overestimate the likelihood of a contextual change (Kaplan et al., 2016), and are more likely to switch between choices in two-choice tasks (Fromm et al., 2023; Schlagenhauf et al., 2014). Computational work implies that these behavioral patterns result from increased beliefs (and uncertainty) about the task volatility (Deserno et al., 2019; Henco et al., 2020; Reed et al., 2020); however, several important issues remain to be resolved.

First, all computational work in this domain only investigated either beliefs about volatility or beliefs about observation noise and has not accounted for both simultaneously and within one common framework. This is an important limitation because the two mechanisms are computationally interdependent, and abnormalities in one might be attributed to abnormalities in the other (Piray & Daw, 2021a). They might also represent two different factors of the illness and recruit different brain circuits (Iglesias, Tomiello, Schneebeli, & Stephan, 2017; Sterzer et al., 2018). Second, findings of increased updating in patients with schizophrenia are hard to reconcile with a range of studies showing patients demonstrate reduced belief updating and impaired flexibility (Baker, Konova, Daw, & Horga, 2019; Doll et al., 2014; Nassar, Waltz, Albrecht, Gold, & Frank, 2021; Waltz & Gold, 2007). Third, a major setback in defining outputs of computational models as clinically relevant phenotypes has been that their test-retest reliability is either unknown (Browning et al., 2020), or modest at best (Enkavi et al., 2019). Despite recent progress demonstrating fair reliability of behavioral patterns in reinforcement learning tasks (Loosen, Seow, & Hauser, 2022; Waltmann, Schlagenhauf, & Deserno, 2022), test-retest scores of parameters representing beliefs about more abstract task features, such as task volatility or outcome variance, are simply not known. Finally, it is not clear to what degree the proposed belief updating patterns are specific to positive symptoms, rather than the syndrome of schizophrenia more generally, and whether they represent transdiagnostic patterns on a whole spectrum of clinical severity.

The present study aimed to address these issues. We first introduced a model that describes how individuals dynamically update their beliefs about environmental volatility as well as observational noise when both can dynamically change throughout the task. We then used the generative computational framework to determine the minimal task conditions under which satisfactory reliability of computational parameters is feasible. We then empirically investigated whether the parameters of the computational model are reliable and clinically meaningful measures. To this end, we first conducted a test-retest study in healthy volunteers and looked at which of the three computational patterns predicts subclinical delusional ideation (Experiment 1). We then replicated and extended the findings of Experiment 1 in a larger data set of healthy participants (Experiment 2), and a data set of patients with schizophrenia (Experiment 3).

## Methods and Materials

### Computational Modelling

The fundamental problem that we need to solve is to provide a principled description of how an agent updates their beliefs about a latent cause in an environment where the (sensory) observations are noisy and the underlying cause changes, whereby this agent’s behavior is determined with a set of specific parameters that could account for interindividual differences. In one simplified example of such an environment, the probabilistic model used to generate outcomes (observations) is a Gaussian variable, where both summary statistics are stable for several trials but can change anytime (Figure 1a). A task with such an outcome generating model has been introduced as the Predictive inference task (Nassar, Wilson, Heasly, & Gold, 2010), whereby one includes reversals in the degree of observational noise within one task session.

**Figure 1.**
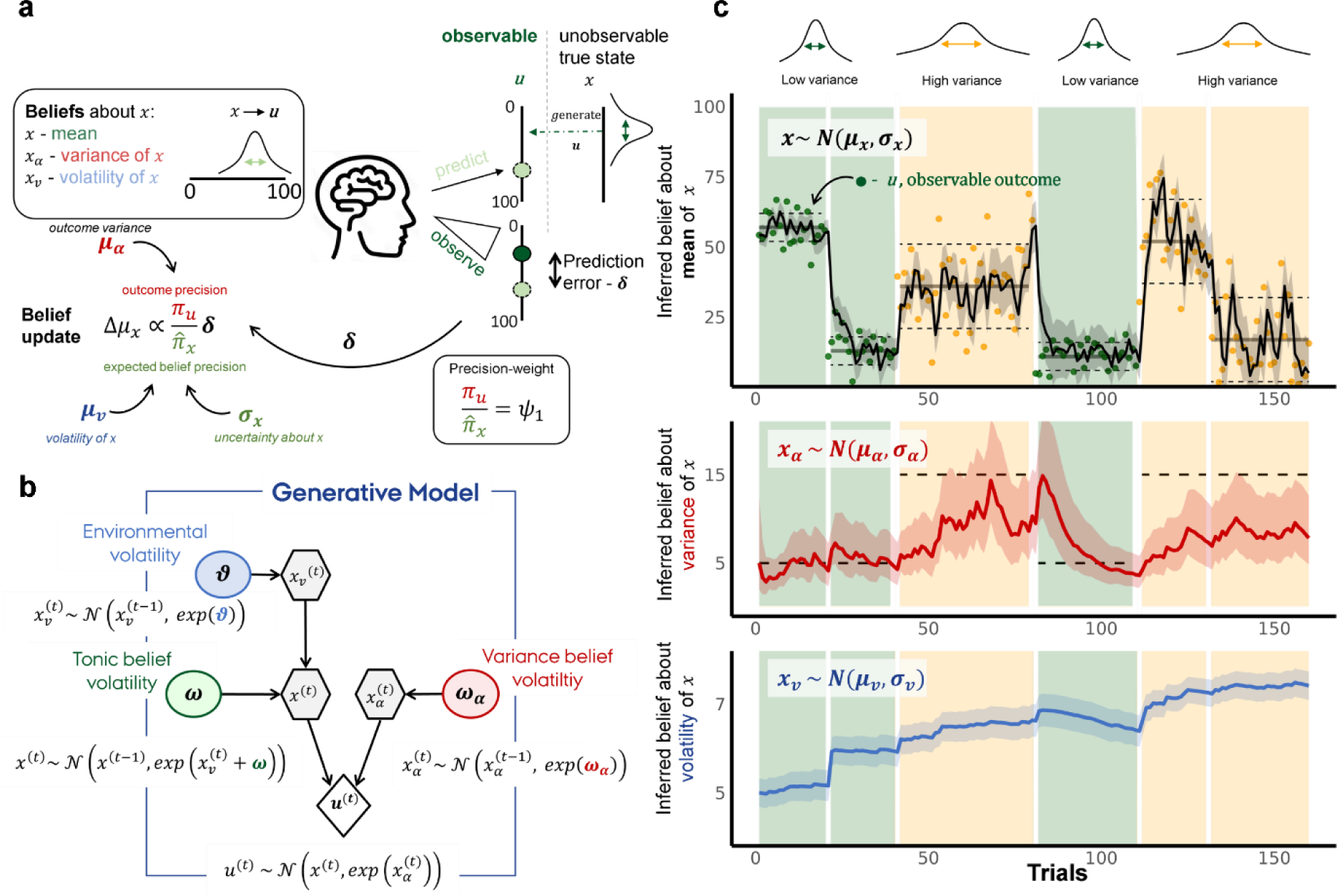
Computational model. **a**, Observations are generated from an unobservable dynamically changing Gaussian variable with varying variance. Observers make predictions based on their beliefs and use the prediction errors to update their beliefs about the underlying states. Belief updates are regulated by the precision weight *ψ*_1_: ratio of the outcome precision (*π*_*u*_) and the expected mean belief precisions (*π*_*x*_), which is in turn governed by both prior belief precision or uncertainty *σ*_*x*_ and belief about volatility *μ*_*v*_. **b**, Generative model of the observer is defined through two cascades of random Gaussian walks that determine the evolutions of beliefs about the mean (and its volatility) and the outcome variance. **c**, Model inversion through the hierarchical Gaussian filter provides both the mean and the precision of beliefs about the mean, variance, and volatility of the latent variable.

The model we present here is based on the generalization of hierarchical generative models invertible by the hierarchical Gaussian filter (HGF, Mathys, Daunizeau, Friston, & Stephan, 2011; Mathys et al., 2014). We define the generative model (Figure 1b) as two parallel cascades of Gaussian random walks, where the first governs the evolution of the underlying mean (*x*), and its (log) volatility (*x*_*v*_), and the second governs the evolution of the outcome variance (*x*_*⍺*_), also on a log scale. The generative model of what causes sensory observations (*u*) is then defined as the mean state *x* and the variance state *x*_*v*_ (Figure 1b). Importantly, the evolutions of the three described states are governed by three agent-specific parameters: variance volatility parameter *ω*_*⍺*_ (the rate of change of the variance state), environmental volatility parameter ʋ (the rate of change of the volatility of the mean), and tonic volatility parameter *ω* (the rate of change of the mean *x*, after accounting for the environmental volatility).

The generative model is inverted using hierarchical Gaussian Filtering (HGF, Mathys, Daunizeau, Friston, & Stephan, 2011; Mathys et al., 2014) to derive at trial-level update equations that resemble other prediction error driven updating, such as reinforcement learning (Sutton, 1992) or the Kalman filter (Gershman, 2015; Kakade & Dayan, 2002). The model inversion is described in detail in work introducing the HGF (Mathys et al., 2011, 2014) and it’s more generalizable form (Weber et al., 2023). Through model inversion we derive inferred posterior distributions of participants’ belief trajectories as Gaussians with the mean *μ* and variance *σ*^2^or its inverse, the precision *π* (see Supplementary Table 1 for update equations). The likelihood function (or the action model) maps beliefs about the mean to behavior (*y*) with a Gaussian function:

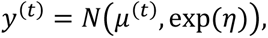

where *η* represents decision noise, or measurement error (on a log scale).

### Parameter estimation

Initial parameter estimation and simulations were done for each individual and session separately using maximum a posteriori estimates and the HGF toolbox in Matlab (Mathworks) and is available at www.translationalneuromodeling.org/tapas as open-source code (GPLv3).

In the test-retest study, we first analyzed behavior without pooling. We compared three different models using the Random-Effects Bayesian Model Selections (Stephan, Penny, Daunizeau, Moran, & Friston, 2009). The priors used for the models are described in Supplementary Table 2.

Because hierarchical inference can improve estimation of parameters across individuals and at the group level (Waltmann et al., 2022), we also analyzed our data within a hierarchical Bayesian framework. Models were implemented in custom code in Stan (Carpenter et al., 2017) using R as the programming language and RStudio as the integrated development environment for R (for the full model definition see Supplementary Methods). Models were estimated using Markov Chain Monte Carlo sampling, with four independent chains and 3000 iterations (1000 warm-up). Convergence of sampling chains was estimated through the Gelman-Rubin *R̂* statistic (Gelman & Rubin, 1992), whereby we considered *R̂* values smaller than or equal to 1.01 as acceptable.

The intraclass correlation (*ICC*) for the parameters was defined as the ratio of between-participant variance and total variance and estimated either with custom code or using the *icc* function from the R package *psych* (using bootstrapping with 1000 samples to obtain distributions).

### Simulating test-retest scores

To calculate test-retest we drew 50 sets of model parameters from the prior distributions (Supplementary Table 2) and set these as unobservable “true” parameters. We generated 2 pairs of task trajectories outcomes (observable inputs) for five different trial numbers (16, 48, 120, 240, and 480). For each of the 50 computational parameter sets we simulate data with three noise levels on two different task versions and estimate the parameters. The re-estimated parameters are then used to calculate the simulated ICC using the *psych* package in R.

### Experiment 1

#### Participants and procedures

Fifty-five (n = 55) healthy adults took part in four in-person testing sessions separated by at least 1 week, out of which n = 45 finished all four sessions and were included in the final analysis. On each session participants performed a behavioral battery that included the Predictive inference task, as well as an exploration-exploitation task and a losing of control task (Mikus, Mancinelli, Lamm, & Mathys, n.d.) that will be published elsewhere. On top of this, in the third session, participants filled out the Peters et al. Delusion Inventory (PDI, Peters, Joseph, Day, & Garety, 2004) and in the fourth session they played the Two-step task (Daw, Niv, & Dayan, 2005; Kool, Cushman, & Gershman, 2016). The participants earned a base fee of € 40 and could earn up to € 80 through bonus payments from the tasks. All participants gave written informed consent, and the experiment was approved by the ethics committee of the University of Vienna (# 1918/2015) and conducted following the latest revision of the Declaration of Helsinki (World Medical Association, 2013).

#### The predictive inference task

For this experiment we adapted the Predictive inference task from Nassar et al. (2010). In this task, participants are required to predict the outcome of each trial. The outcome is a random number between 0 and 100, drawn from a normal Gaussian distribution with a certain mean and standard deviation, with a rebounding boundary. In contrast to the task by Nassar and colleagues, both the mean and the standard deviation change several times throughout the task. The probability of the mean changing is 0.1 except for the first 3 trials of each block (as in Nassar et al. 2010, 2012) and the probability of the standard deviation to change when the mean changed is 0.4, and could be either high (15) or low (5). Participants could express their confidence with a longer button press and performance was incentivized based on prediction errors. Participants played 240 trials in four sessions with four different trajectories that did not change across participants. For a detailed description of the task see Supplementary Methods.

#### The two-step task

The two-step task is often used as a behavioral measure for the capacity for model-based decision making (Daw et al., 2005; Kool et al., 2016), whereby keeping a “model” of the mapping between step one and step two can improve points earned in the task. For a detailed description of the task refer to the Supplementary Methods. For 5 participants, the data of the task was not saved, therefore, a total of 40 participants were included in the analysis of this data.

#### Behavioral analysis

For frequentist mixed linear models, the lme4 package in R was used, and for Bayesian mixed linear models, the brms package was used. For frequentist models we report Confidence Intervals (CI)s and p values (P). for Bayesian models we report Credibility intervals (CrI) and the probability of the posterior interval that lies above (or below) zero (P). For details on behavioral analysis see Supplementary Methods.

### Experiment 2

In the second experiment, we reanalyzed data from a previously published online study (for details see Seow & Gillan, 2020). Here, a sample of 437 (249 female) MTurk participants, aged 20–65 (mean = 36.3. SD = 10.2) played a version of the Predictive inference task with constant noise and varying volatility (see Supplementary Methods for details) and filled out a wide range of questionnaires assessing general psychopathology including the Short Scales for Measuring Schizotypy that consists of four subscales (Mason, Linney, & Claridge, 2005). As described in Seow and Gillan (2020), all questionnaires together consisted of 209 items, and weights from a factor analysis done in a previous study (Gillan, Kosinski, Whelan, Phelps, & Daw, 2016) were used to define three factor scores named as “compulsive behavior and intrusive thought” (CIT), “anxious-depression” (AD), and “social withdrawal” (SW).

### Experiment 3

In the third experiment, we reanalyzed data from a previously published study with patients with schizophrenia (Nassar et al., 2021). Data was used from 108 participants with a diagnosis of schizophrenia or schizoaffective disorder collected at the Maryland Psychiatric Research Center, University of Maryland School of Medicine. Participants were stable outpatients, most treated with antipsychotic medications. Inclusion criteria in patients were a presence of a schizophrenia spectrum disorder in patients. Exclusion criteria were a presence of a current Axis I disorder, a neurological disorder, or a cognitively impairing medical disorder. The severity of positive and negative symptoms were assessed with the Brief Psychiatric Rating Scale (BPRS, (Overall & Gorham, 1962)), and Scale for the Assessment of Negative Symptoms (SANS, (Andreasen, 1989)), respectively. In the predictive inference task performance was incentivized in two different ways across two different task sessions, each lasting for 100 trials. In the appetitive condition, participants could earn points and in the aversive condition, participants would have points taken away from their initial endowment in a performance contingent way (see Supplementary Methods for details).

## Results

### Computational Model

The three model parameters *ω*, *ω*_*⍺*_ and ʋ determine interindividual (and inter-session) differences in how participants update beliefs about the mean (*μ*), the outcome variance (*μ*_*⍺*_) or the task volatility (*μ*_*v*_), by specifying the precision weights on the respective belief trajectories (see update equations Supplementary Table 1). Low values of *ω*, *ω*_*⍺*_ and ʋ imply rigid beliefs about the mean, the outcome variance or environmental volatility, respectively (see Supplementary Figure 1 for model simulations with various combinations of parameter sets).

To make sure the three parameters correspond to distinct behavioral patterns we looked at parameter recovery and found high correlations between simulated and retrieved parameters under low noise assumption, with 480 trials (r>0.96). However, as decision noise increases, or as trial numbers decrease, parameter recovery becomes harder (Supplementary Figure 2).

Next, we aimed to determine under which conditions the reliability of computational parameters in an experiment would be sufficient. In other words, when can a task generating process (a jumping Gaussian in our case), coupled with a model, produce computational parameters that show good intra-participant reliability across testing sessions (Figure 2a). ICCs were dependent on trial numbers and decision noise and were generally high for the tonic belief volatility parameter and more varied for the environmental and variance volatilities. Yet, with sufficient trials and low decision noise, parameters are retrievable even across tasks with different trial numbers (Supplementary Figure 3). In contrast, when looking at simulated ICC scores for binary data with binary outcomes, using a binary version of the HGF often used modelling behavior in reversal learning tasks, we found that even with high trial numbers and low noise, environmental volatility does not have satisfactory ICC scores (Figure 2b).

**Figure 2.**
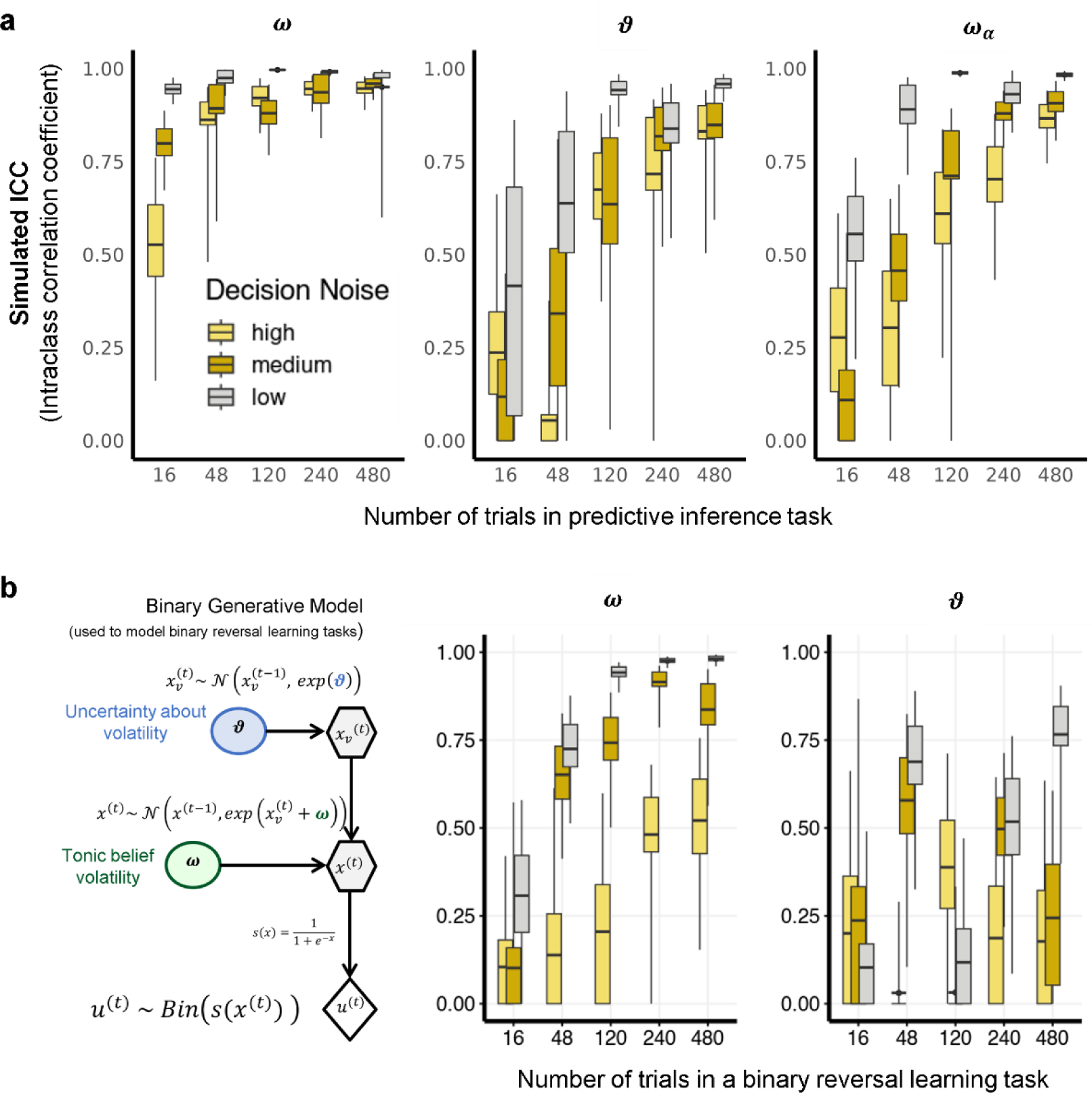
Simulated intraclass correlations. **a**, Continuous predictive inference task simulation with decision noise *η* (1, 16, or 48). **b**, Binary reversal one-armed bandit task, where the trajectories (correct/incorrect) are generated with the same probability of reversal as in (a) and the probability of correct in each block being either 0.2, 0.8, or 0.5. The decision noise is parametrized through the softmax inverse temperature set as 48, 8, or 1 (from low to high noise).

### Experiment 1

#### Model and behavior

We first wanted to make sure that participants in our task tend to adapt their learning rate according to higher variances in outcomes. As above, we define the learning rate as the ratio between the prediction update and the prediction error. We found that participants on average reduced their learning rate in high compared to low outcome variance blocks (b =-0.167, SE = 0.011, d= 0.168, t(139.34) = −15.761, P<0.001, Figure 3a). We compared the model with all three parameters to models that would (correctly) assume that the environmental volatility does not change. We found that the model with all three parameters outperformed the other two models in all sessions (Supplementary Table 3), suggesting that on average participants tended to adjust their beliefs about the volatility of the mean throughout the task. In line with this, we found that the (model implied) mean beliefs about volatility were higher for high compared to low outcome variance blocks (b = 0.108, SE = 0.015, d = 0.190, t(55.245) = 7.199, P < 0.001, Figure 3b), as were mean beliefs about noise (b = 0.14519, SE = 0.01588, d = 0.265, t(52.504) = 9.141, P<0.001, Figure 3c).

**Figure 3.**
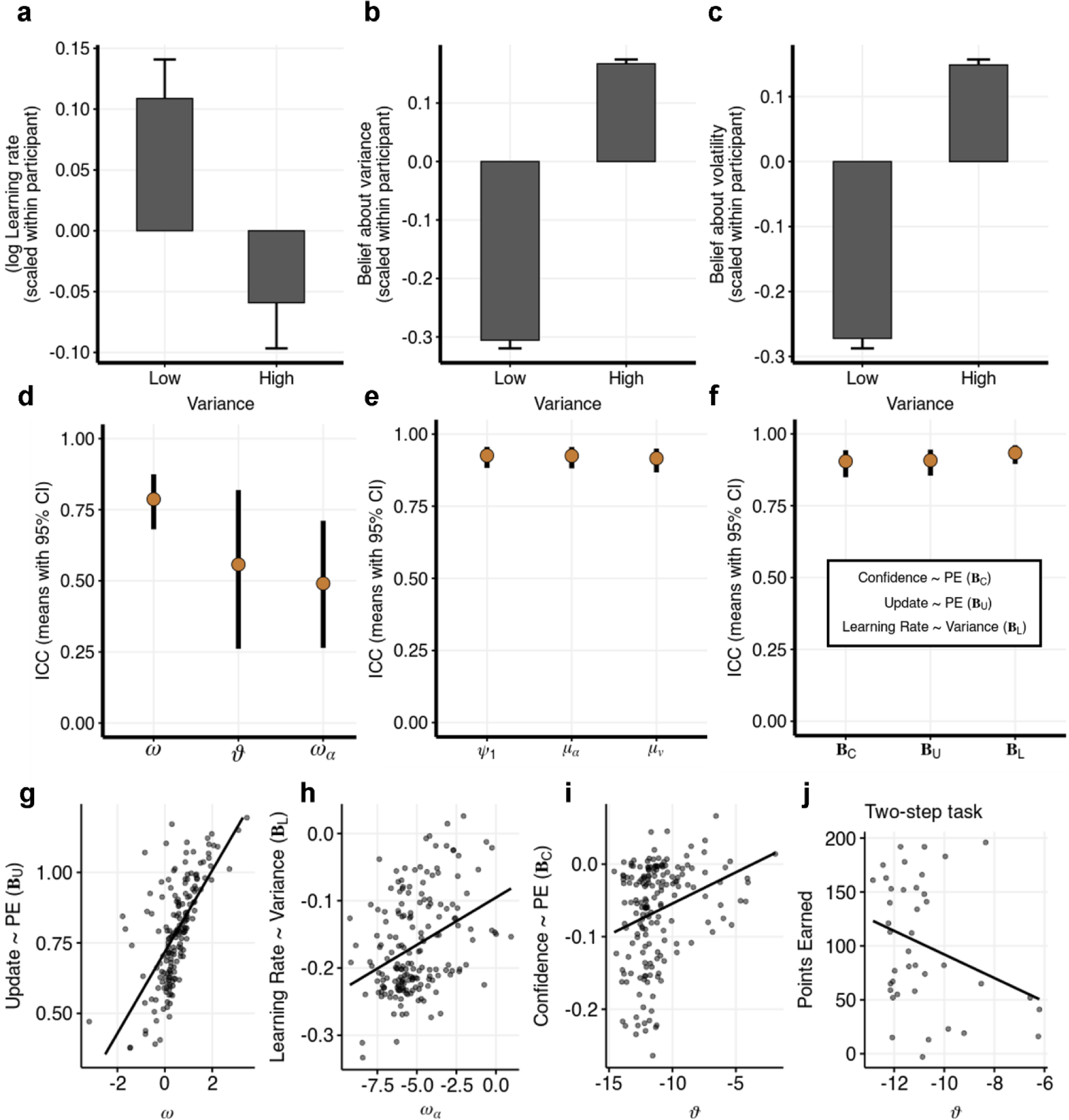
Model performance in Experiment 1. **a – c**, Learning rate adaptation (a), beliefs about variance (b) and volatility (c) across the variance blocks. All dependent variables were scaled within participant, then averaged within participant and variance block and then across participants. Bar plots and error bars represent means and standard errors across participants (with N = 45). **d**, Empirical ICC scores derived from the hierarchical model (means and 95% CrI). **e**, ICCs (means and 95% CI) of model derived trial-by-trial precision-weighted learning rates (*ψ*_1_), beliefs about outcome variance (*μ*_*⍺*_) and volatility (*μ*_*v*_) meaned within participant. **f**, ICCs (means and 95% CI) plotted for the participant-level random slope of prediction error effect on confidence on the next trial (***B***_*C*_), singed prediction error on signed update (***B***_*U*_), and the effects of variance on learning rate (***B***_*L*_). **g – i**, Relationship of random slopes ***B***_*U*_, ***B***_*L*_, and ***B***_*C*_ on model parameters. **j**, Correlation between the performance in the Two-step task and the environmental volatility parameter ʋ in session 1 (N = 40).

#### Test-retest

ICC scores (derived from the hierarchical models as ratios of posterior distributions of between-participant variance and total variance) were the following: for *ω* the mean was 0.787 (95% CrI [0.681, 0.874]), for ʋ the mean was 0.557 (95% CrI [0.261, 0.819]) and for *ω*_*⍺*_ the mean was 0.491 (95% CrI [0.264, 0.711], Figure 3d), indicating at least moderate ICCs. However, when looking at the ICC scores of averaged trial-by-trial trajectories calculated with a separate linear model (Figure 3d), we found excellent test-retest scores for the precision-weighted learning rates *ψ*_1_(ICC = 0.93, 95% CI [0.88, 0.96], for mean beliefs about volatility *μ*_*v*_ (ICC = 0.92, 95% CI [0.87, 0.95]), as well as noise *μ*_*⍺*_ (ICC = 0.92, 95% CrI [0.88, 0.96]). The high ICCs are likely because hierarchical estimation pulls estimates within participants closer together. Note that the ICC scores are lower (in particular for the *μ*_*v*_) when parameter estimation was done without pooling, or when looking at only two sessions at a time (Supplementary Figures 4, 5 and 6).

In the next step, we looked at how behavior in the task related to model parameters, and how stable it was across sessions. We defined three participant-level behavioral patterns as participant-level regression slopes of 1) the model predicting updates on the next trial from signed prediction error (***B***_***U***_); 2) the model estimating how the behavioral learning rate is affected by outcome variance (***B***_***L***_); and 3) the model predicting confidence ratings on the next trial from absolute (unsigned) prediction errors (***B***_***C***_). We found that the all three participant-level random slopes have excellent ICC (all mean ICC > 0.9, Figure 3e). Looking at how the three model parameters predict each of the three behavioral patterns, we found *B*_*U*_ was most strongly predicted by the belief volatility parameter *ω* (b = 0.795, SE = 0.053, t = 14.884, P < 0.001, Figure 3g, Supplementary Table 4), *B*_*L*_ correlated only with the variance belief volatility parameter *ω*_*⍺*_ (b = 0.438, SE= 0.115, t = 3.822, P < 0.001, Figure 3h, Supplementary Table 5), and *B*_*C*_ correlated ʋ (b = 0.356, SE = 0.111, t = 3.202, P = 0.002 r = 0.280, t(178) = 3.888, P < 0.001, Figure 3i, Supplementary Table 6).

The environmental volatility parameter represents the amount of uncertainty participants have about how volatile the environment is and is conceptually related to the ability to form and maintain high-order models about the world. This is comparable to the concept of model-based learning in the reinforcement learning framework. In support of this, we found some evidence that the performance of participants in the two-step task (points earned) correlated negatively to ʋ (*r*_*session*1_ = −0.371, P = 0.048, *r*_*session*2_ = −0.268, P = 0.097, *r*_*session*3_ = −0. 317, P = 0.049, *r*_*session*4_ = - 0.226, P = 0.101, N=40 Figure 3j), but not with *ω*_*⍺*_ (all P >0.4), nor *ω* (all P > 0.16).

#### Schizotypal traits correlate with the environmental volatility parameter

When looking at how the mean parameters across all four sessions predict PDI scores (Figure 4 a)we found that higher PDI was related to increased environmental volatility parameter ʋ (effect size d = 0.687, 95% CrI [0.027, 1.373], P(d<0) = 0.02, Figure 4 b), and no evidence for a decrease in the noise volatility parameter *ω*_*⍺*_ (d = −0.548, 95% CrI [−1.317, 0.198], P(d>0) = 0.072) nor on the tonic volatility parameter *ω* (d = 0.176, 95% CrI [−0.407, 0.771], P(d<0) = 0.286, for session-by-session analysis, and results of the analysis with parameters estimated without pooling, see Supplementary Table 7).

**Figure 4.**
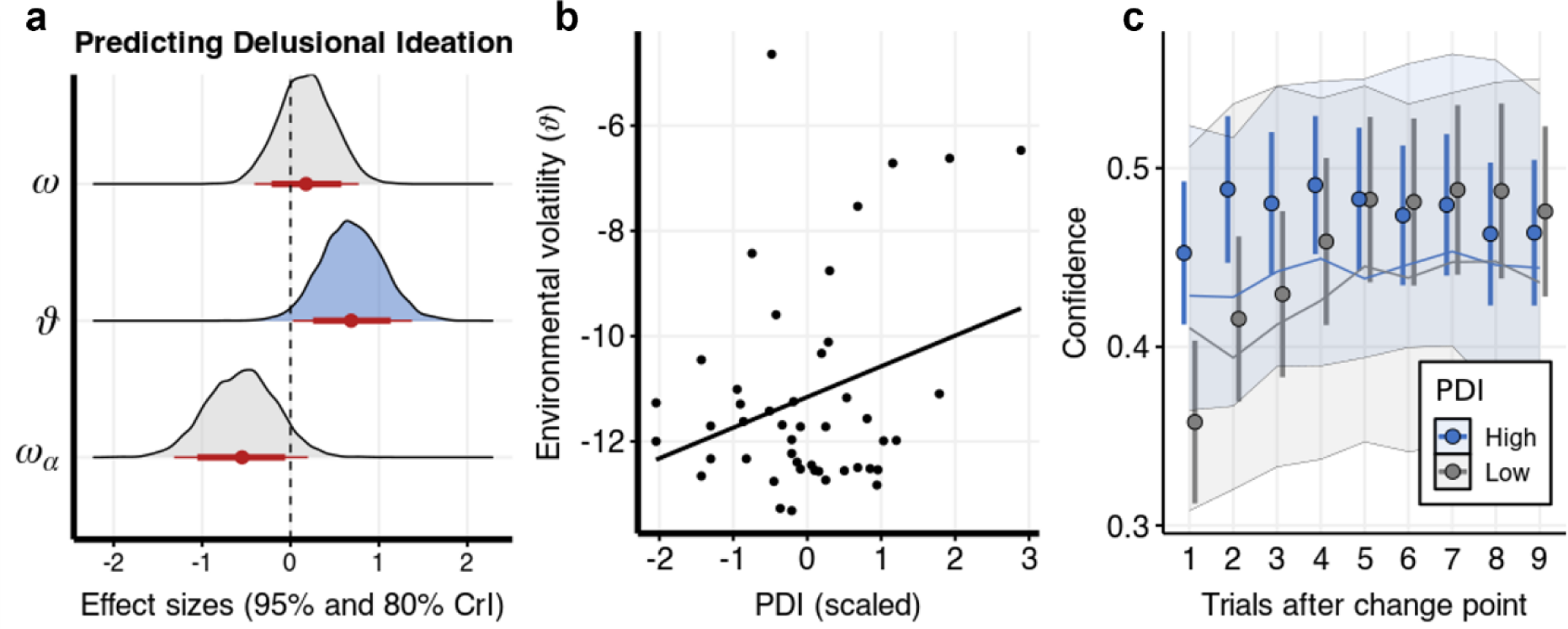
Model parameters and schizotypal traits. **a**, Posterior distributions of effect sizes of the effect of each model parameter (meaned per participant) predicting PDI, over means and CrIs. **b,** Correlation plot of ʋ and PDI. PDI was log transformed and scaled. **c,** Plotting confidence ratings after change points. Points and error bars represent means and standard errors (across participants). Line and shade represent the posterior predictive mean and 95% Predictive interval of a linear regression model predicting confidence from prediction errors. PDI: Peters et al. Delusional Inventory.

We then investigated how these effects are mirrored in the behavioral patterns described above. Based on the results from the previous section we expected that PDI scores would decrease the effect of prediction errors on the confidence rating in the next trial (*B*_*C*_). However, we found no robust evidence of PDI’s effect on confidence behavior (*b*_*PE*:*PDI*_ = 0.009, 95% CrI [−0.011, 0.029], P(b<0) = 0.186). Plotting the posterior predictions of the model we see that the model poorly captures confidence ratings after change points with a high uncertainty in predictions (Figure 4 c).

### Experiment 2

We analyzed the data from (Seow & Gillan, 2020) with the same computational model as in Experiment 1 and investigated how the model parameters relate to schizotypal traits. Predicting the total scores of the Short Scale for Measuring Schizotypy from model parameters, controlling for age, gender, and IQ we found positive correlations with the environmental volatility parameter ʋ (d = 0.14, 95% CrI [−0.007, 0.287], P(b<0) = 0.032, Supplementary Figure 8a), but not with *ω*_*⍺*_ (P(d>0) = 0.45), nor *ω* (P(d>0) = 0.171). We then further explored how our parameters predict two subscales of schizotypal traits: “Unusual Experiences” (UE) and “Introvertive Anhedonia” (IA, see results for the other two subscales in the Supplementary Figure 8b). Predicting the scores in the UE subscale (Figure 5a, Supplementary Table 8) we found positive correlations with the environmental volatility parameter ʋ (d = 0.092, 95% CrI [−0.001, 0.186], P(d<0) = 0.026), but not with *ω*_*⍺*_ (d = −0.013, 95% CrI [−0.109, 0.086], P(d>0) = 0.401), nor *ω* (d = −0.045, 95% CrI [−0.14, 0.051], P(d>0) = 0.171). In contrast, the IA subscale (Figure 5b, Supplementary Table 9) did not correlate reliably with ʋ (d = 0.029, 95% CrI [−0.066, 0.123], P(d<0) = 0.286), but we found weak evidence of a negative correlation with *ω*_*⍺*_ (d = −0.081, 95% CrI [−0.179, 0.017], P(d>0) = 0.053) and *ω* (d = −0.071, 95% CrI [−0.168, 0.023], P(d>0) = 0.071). When investigating how both subscales modulated the effect of prediction error on confidence ratings on the next trial (Figure 5c, Supplementary Table 10), we found a significant interaction effect of UE (d = 0.035, 95% CrI [0.015, 0.055], P(d<0) < 0.001), but not IA (d = 0.01, 95% CrI [−0.009, 0.029], P(d<0) = 0.154). Interestingly, we also found that higher UE scores predicted higher confidence ratings on average (d = 0.151, 95% CrI [0.102, 0.198], P(d<0) = 0.001), whereas IA scores predicted lower confidence on average (b = −0.051, 95% CrI [−0.096, - 0.001], P(b>0) = 0.023).

**Figure 5.**
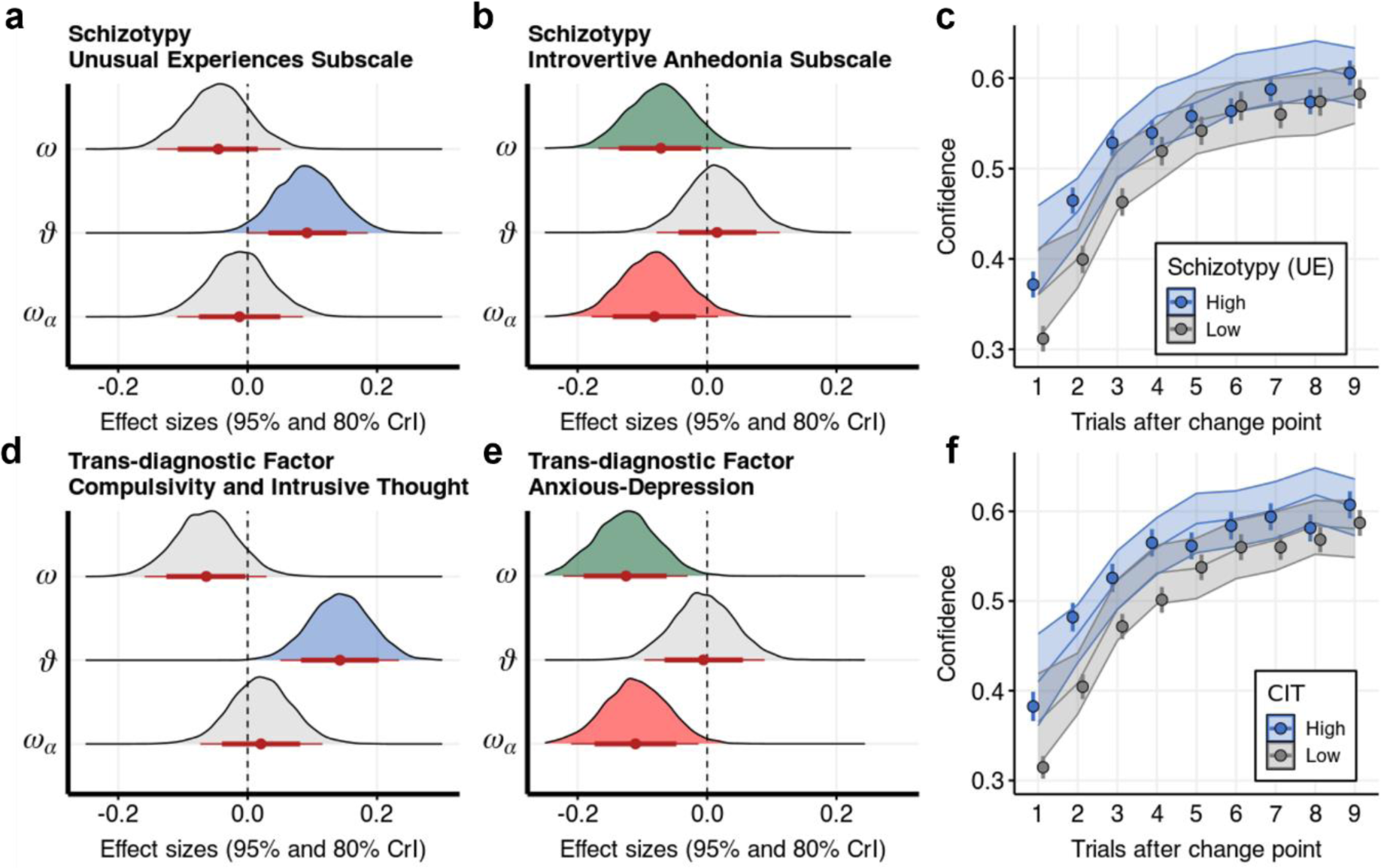
Modelling results from an online sample of healthy participants. **a-b**, Two subscales of the Short Scale for measuring Schizotypy were predicted by the three parameters controlling for age, gender and IQ. Plotting posterior distributions of effects of each model parameter, over means and CrIs of effect sizes. **c,** Plotting confidence ratings after change points for the median split of the Unusual Experiences scale. Points and error bars represent means and standard errors (across participants). Line and shade represent the posterior predictive mean and 95% Predictive interval from a linear regression model predicting confidence from prediction errors. **d-e**, CIT and AD predicted by model parameters controlling for age, gender and IQ in two separate models. Plotting posterior distributions of effects of each model parameter over means and CrIs of effect sizes. **f,** Plotting confidence ratings after change points. Points and error bars represent means and standard errors (across participants). Line and shade represent the posterior predictive mean and 95% Predictive interval of a linear regression model predicting confidence from prediction errors.

We then investigated to what degree these effects are specific to traits related to schizotypy. We looked into how our model parameters predicted the first two factors of the factor analysis, named as “compulsive behavior and intrusive thought” (CIT), and “anxious-depression” (AD). In a model that predicted the CIT factor from model parameters, controlling for age, gender and IQ, we found a positive effect of the ʋ parameter (d = 0.143, 95% CrI [0.05, 0.234], P(d<0) = 0.001, Figure 5d, Supplementary Table 11), but not *ω* (d = --0.064, 95% CrI [−0.159, 0.029], P(d>0) = 0.088), nor *ω*_*⍺*_ (d = 0.021, 95% CrI [−0.073, 0.116], P(d<0) = 0.331). In contrast, the AD factor was negatively related to *ω* (d = −0.125, 95% CrI [−0.222, −0.03], P(d>0) = 0.006, Figure 5e, Supplementary Table 12), as well as *ω*_*⍺*_ (d = −0.111, 95% CrI [−0.21, −0.013], P(d>0) = 0.013), but not ʋ (d = 0.006, 95% CrI [−0.097, 0.089], P(d<0) = 0.457).

We also found that the factors modulated the effect of prediction error on confidence ratings on the next trial (Figure 5f, Supplementary Table 13), with CIT increasing confidence on average (d = 0.22, 95% CrI [0.168, 0.271], P(d<0) < 0.001), as well as having a positive interaction with the PE (d = 0.058, 95% CrI [0.037, 0.08], P(d<0) < 0.001), and AD decreasing confidence on average (d = −0.098, 95% CrI [−0.154, −0.045], P(d>0) < 0.001), but did not modulate the effect of the prediction error (d = 0.008, 95% CrI [−0.015, 0.032], P(d<0) = 0.256).

### Experiment 3

To see to what extent these results extend to the clinical domain of positive and negative symptoms of schizophrenia we analyzed the behavior of 108 patients with schizophrenia (previously published in Nassar et al., 2021). The behavior in the task was analyzed with the same model as in Experiments 1 and 2, whereby we estimated the parameters for the two incentivization conditions (reward seeking and loss avoiding) separately. We first looked at how well the parameters of the model predict positive symptoms, controlling for age, gender, and IQ. Because patients might display a high degree of noise in their behavior, we also included decision noise as a predictor in the models.

Using separate parameters for reward seeking and loss avoiding conditions, we found that the environmental volatility parameter ʋ estimated in the reward seeking condition was a significant predictor of positive symptoms (d = 0.187, 95% CrI [−0.019, 0.403], P(d<0) = 0.039, Figure 6a, Supplementary Table 14), but we found no credible support for *ω* (d = −0.082, 95% CrI [−0.301, 0.137], P(d>0) = 0.235) nor *ω*_*⍺*_ (b = −0.168, 95% CrI [−0.392, 0.057], P(b>0) = 0.07). We also found no evidence for an association between the parameters estimated in the loss avoidance condition on positive symptoms (all P>0.326). When predicting negative symptoms, we found that in the loss avoiding condition, there was a negative association between *ω*_*⍺*_ (d = −0.298, 95% CrI [−0.523, - 0.088], P(d>0) = 0.003, Figure 6b, Supplementary Table 15) and *ω* (d = −0.223, 95% CrI [−0.442, 0.003], P(d>0) = 0.026), but not ʋ (d = −0.012, 95% CrI [−0.238, 0.216], P(d>0) = 0.457). We found no evidence for an association of negative symptoms with the model parameters in the reward seeking condition (all P > 0.214).

**Figure 6.**
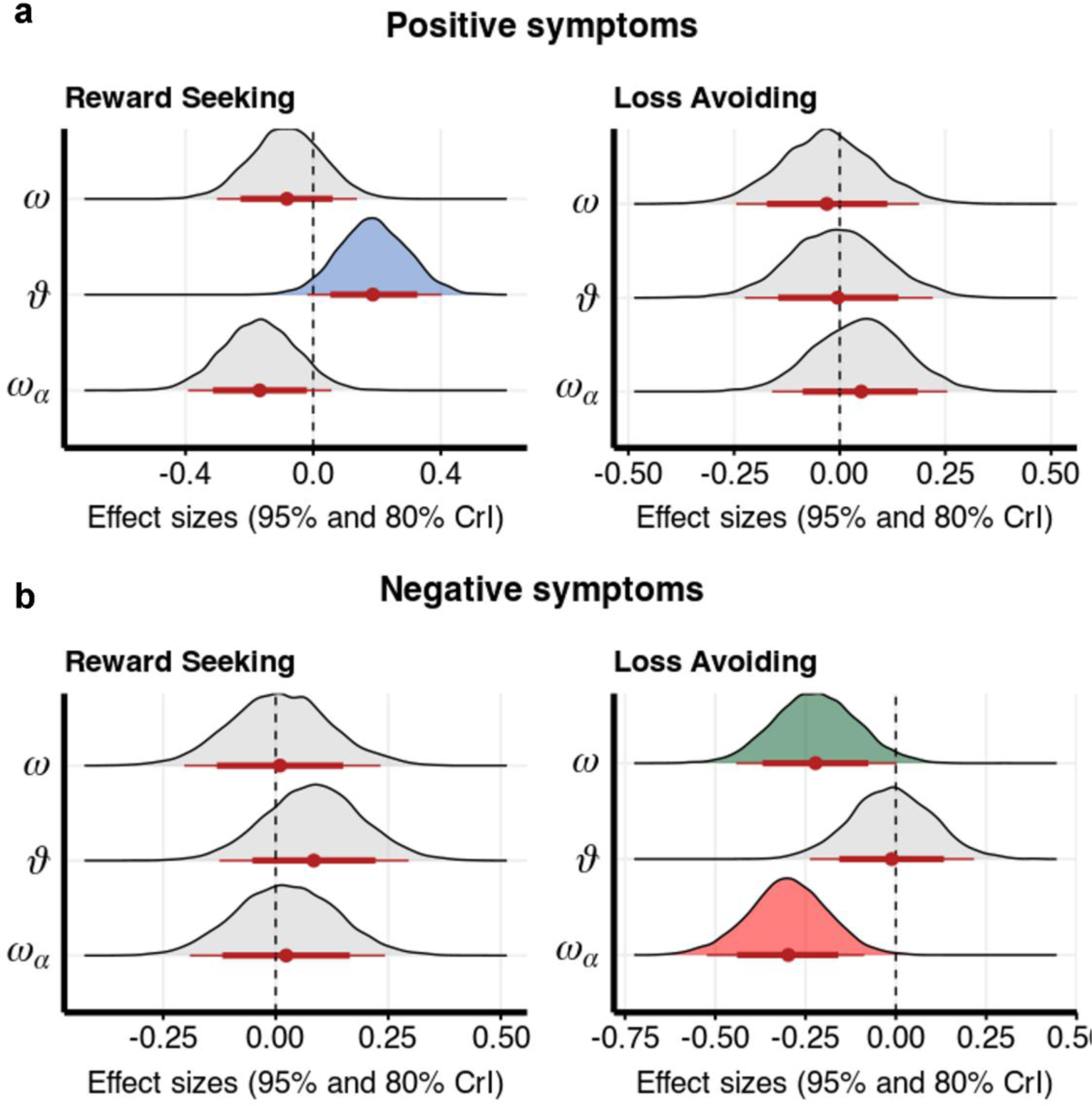
Modelling results from a sample of patients with chronic schizophrenia in Experiment 3. A total of eight parameters (three parameters of the belief model as well as a noise parameter *η* (on a log scale) for both reward seeking and loss avoiding condition) and age, gender, and IQ were used as predictor variables in two models predicting Positive symptoms measured by the Brief Psychiatric Rating Scale (**a**) and negative symptoms measured by the Scale for the Assessment of Negative Symptoms (**b**). Plotting posterior distributions of effects of each model parameter, over means and CrIs of effect sizes across two incentivization condition.

## Discussion

In this paper, we present a computational model that describes how individuals with schizophrenia and schizotypal traits learn in environments where both observational noise and the volatility of environmental events can change. We demonstrate that schizotypal traits and positive symptoms in schizophrenia were related to higher uncertainty about the task volatility, while depressive-anxious traits and negative symptoms were associated with rigid beliefs about the underlying mean and outcome variance.

Previous work does suggest that schizotypal traits correlate with volatility estimates across both clinical and nonclinical samples (Deserno et al., 2019; Reed et al., 2020), however, not accounting for participants’ perception of outcome reliability limits the conclusions that can be drawn from these studies (Piray & Daw, 2021b; Pulcu & Browning, 2019). In tasks with binary outcome variables, outcome variance is determined from the probability of outcome and therefore indistinguishable from prior beliefs. Importantly, even in stable task environments (without latent changes), patients with schizophrenia do not down-weight their updating in response to higher outcome variance (Haarsma et al., 2020) and there are several known examples of patients failing to attenuate sensory signals (Adams et al., 2013; Schmack, Schnack, Priller, & Sterzer, 2015; Sterzer et al., 2018; Weilnhammer et al., 2020). According to our results, in tasks with latent changes, positive symptoms in chronic patients and schizotypal trats are related to an inability or unwillingness to extract and utilize higher-order statistical features of the environment – processes that are encoded at higher levels of neural computation such as the hippocampus and the prefrontal cortex (Heinz et al., 2019; Lisman & Grace, 2005; Sterzer et al., 2018).

In support of this disrupted “model” learning or utilization, we found some evidence that the uncertainty about the volatility parameter is related to reduced performance in the Two-step task, a task commonly used to assess model-based behavior and prospective decision-making. These results link behavior in reversal learning tasks of individuals with schizotypal traits or positive symptoms to previous work showing impaired model-based control (Culbreth, Westbrook, Daw, Botvinick, & Barch, 2016), or reduced goal-directed behavior and cognitive control in patients more generally (Cohen, 1996; Collins, Albrecht, Waltz, Gold, & Frank, 2017; Gold, Waltz, & Frank, 2015).

It is worth noting however, that higher uncertainty about the task volatility can have adaptive value since it allows for a rapid reduction of uncertainty and higher confidence when faced with unpredictability (Johnson & Fowler, 2011). In our data, higher confidence despite poor predictions was related to higher volatility uncertainty, as well as to schizotypal traits, in line with findings that patients with schizophrenia display overconfidence across a range of social, cognitive and learning tasks (Hahn, Moritz, Elmers, & Scheunemann, 2021; Hoven et al., 2019; Köther et al., 2012; Moritz et al., 2014; Rossi-Goldthorpe, Leong, Leptourgos, & Corlett, 2021). Modelling work has shown how having a wide prior over alternative hypotheses can serve as a “protective belt” when faced with surprising outcomes (Gershman, 2018), which allows for explaining away unpredictable events with a new hypothesis (e.g., a new change has occurred), while being in line with the former belief being held with high confidence (Erdmann & Mathys, 2022). Within this framework, patients can display reduced belief updating (Waltz & Gold, 2007) and a bias against confirmatory evidence (Adams, Napier, Roiser, Mathys, & Gilleen, 2018; Doll et al., 2014), despite displaying sudden shifts in beliefs (Nassar et al., 2021), thereby reconciling the rigidness and confidence with which delusional beliefs are held, and the little evidence necessary to form those beliefs.

Our finding that rigid beliefs about outcome and outcome variance are related to the anxious-depression transdiagnostic psychiatric dimension is an important addition to previous modelling and empirical work (Pulcu & Browning, 2019). In a binary reversal learning task where shocks are paired with stimuli, one study showed that participants with higher trait anxiety were shown to less strongly adjust their learning rate (and pupillary response) to task volatility changes (Browning, Behrens, Jocham, O’Reilly, & Bishop, 2015) and another found a positive correlation between chronic stress and uncertainty about task volatility (De Berker et al., 2016). Recent modelling work with a joint model for outcome variance (stochasticity) and volatility suggests that the relationship between anxiety and learning rate adaptations is best explained by the degree to which surprising events are attributed to volatility vs variance belief updates (Piray & Daw, 2021a). Our results provide empirical support for this claim, whereby individuals high on the anxious-depression dimension had more rigid beliefs about outcome variance, suggesting they were more likely to attribute surprising events to environmental changes. Similarly, we found the same pattern of reduced learning about variance and over all reduced learning being related to negative symptoms of patients in schizophrenia, but only when learning to avoid losses with no credible evidence for an effect when learning about gains. This is in accordance with the idea that negative symptoms arise from a reduced neural response to surprising events (Deserno, Heinz, & Schlagenhauf, 2015; Maia & Frank, 2017), but seems to be at odds with a range of studies showing that this pattern is specific for reward and not avoidance learning (Gold et al., 2013, 2015; Strauss et al., 2011). This discrepancy could be due to most studies using binary choice outcomes, where loss avoidance is conceptualized as absence of action. This contrasts with our study, where behavior of participants reflects their inference about the underlying cause.

One important contribution of this study is the conceptual framework within which a task and model pair can be optimized for good test-retest reliability scores. We showed with simulations that intra class coefficients of parameters can sometimes be improved with higher trial numbers, and that, in some cases, it not feasible to expect high parameter reliability, even with high trial numbers, such as estimating environmental volatility parameters using binary outcomes and behavioral inputs. This has three relevant implications. First, for reliable intra session measurement, tasks where participants learn about continuous input are preferred over binary inputs; second, simulations can be used to optimize the task generating process (for example, by restricting trajectories); and third, model selection can be contingent on the test-retest reliability of the computational parameters given the data. In line with previous work (Zech et al., 2022), we showed that empirical ICCs are improved with hierarchical estimation, which suggests that more sessions per individual will give more precise ICC estimates, as would standardization of experimental designs and models and extending to response models to include several data sources (e.g. confidence, or physiological data).

Despite the consistency of our main findings across the three studies, the effect sizes are low in the two higher powered studies (Experiment 2 and 3). Meta-analytic evidence indicates that this is usually the case when considering specific associations of belief updating patterns with positive symptoms (Dudley, Taylor, Wickham, & Hutton, 2016; McLean, Mattiske, & Balzan, 2017). These measures can nevertheless have a clinically meaningful impact (Funder & Ozer, 2019; Meyer et al., 2001), especially considering the possibility of repeated measurement that current standardized questionnaires are not suited for. However, to explain the heterogeneity of experiences across the psychosis continuum several other processes beyond belief updating (at least as they are operationalized within such experimental tasks) will have to be considered, such as the subjective nature, the content and the context of such experiences (Feyaerts, Henriksen, Vanheule, Myin-Germeys, & Sass, 2021).

In conclusion, we show that mechanistic descriptions of how individuals with psychotic symptoms and psychotic-like traits process information when faced with various sources of uncertainty have the potential to become reliable, transdiagnostic and clinically meaningful computational phenotypes. In future work, the robustness and reliability of these measures can be improved through larger standardization studies and extending response models to other data modalities. Clinical usability, such as predictive validity, can be explored by embedding the computational phenotypes within interventional clinical designs.

## Data Availability

Data will be available online (https://github.com/nacemikus/jget-schizotypy.git) after the peer-reviewed publication of the manuscript.

https://github.com/nacemikus/jget-schizotypy.git

