## Supplementary Material for "Computational phenotyping of aberrant belief updating in individuals with schizotypal traits and schizophrenia"

Nace Mikus [1,2], Claus Lamm[1,*] & Christoph Mathys[2,3,4,*]

1 Department of Cognition, Emotion, and Methods in Psychology, Faculty of Psychology, University of Vienna, Austria;

2 Interacting Minds Centre, Aarhus University; Denmark

3 Translational Neuromodeling Unit, University of Zurich and ETH Zurich, Zurich,

Switzerland;

4 Scuola Internazionale Superiore di Studi Avanzati (SISSA), Trieste, Italy

*Shared senior authors

Nace Mikus1,2, Claus Lamm1* & Chris Mathys2,3,4*

#### Supplementary Tables

**Supplementary Table 1.** Hierarchical Gaussian Filtering of the generative model returns inferred beliefs with update equations for each agent, given the free parameters $\omega$, $\omega_\alpha$, and $\vartheta$.

| Beliefs | Inferred beliefs | Update equations |
|---|---|---|
| Mean | $x^{(t)} \sim N(\mu^{(t)}, \sigma^{(t)})$ | $\Delta\sigma = \exp\left(\mu_v^{(t)} + \boldsymbol{\omega}\right)$<br>$\pi^{(t)} = 1/\sigma^{(t)}$<br>$\hat{\pi}^{(t)} = \pi^{(t)} + \pi_u^{(t)}$<br>$\pi_u^{(t)} = 1/\exp\,(\mu_\alpha^{(t)})$<br>$\psi_1 = \frac{\pi_u^{(t)}}{\hat{\pi}^{(t)}}$<br>$\Delta\mu = \psi_1\delta$ |
| Volatility | $x_v^{(t)} \sim N\left(\mu_v^{(t)}, \sigma_v^{(t)}\right)$ | $\Delta\sigma_v = \exp(\boldsymbol{\vartheta})$<br>$\pi_v = 1/\sigma_v$<br>$\Delta\mu_v \propto \frac{1}{\pi_v}\ \delta_v$ |
| Outcome variance | $x_\alpha^{(t)} \sim N\left(\mu_\alpha^{(t)}, \sigma_\alpha^{(t)}\right)$ | $\Delta\sigma_\alpha = \exp\,(\boldsymbol{\omega_\alpha})$<br>$\pi_\alpha = 1/\sigma_\alpha$<br>$\Delta\mu_\alpha \propto \frac{1}{\pi_\alpha}\ \delta_\alpha$ |

**Supplementary Table 2.** Priors for the three HGF models used to analyze data from the Test-retest study (N=45).

| | HGF 1 | HGF 2 | HGF 3 |
|---|---|---|---|
| | Model with all uncertainty parameters, but fixed initial variance and volatility belief | Model with a fixed environmental volatility parameter, but free initial volatility estimate | Model with a fixed environmental volatility parameter, and fixed initial variance and volatility belief |
| Priors | $\omega \sim N(-3,5)$<br>$\omega_\alpha \sim N(-3,5)$<br>$\vartheta \sim N(-3,5)$<br>$\mu_v^{(0)} = 4$<br>$\mu_\alpha^{(0)} = 5$ | $\omega \sim N(-3,5)$<br>$\omega_\alpha \sim N(-3,5)$<br>$\exp(\vartheta) = 0$<br>$\mu_v^{(0)} \sim N(4,5)$<br>$\mu_\alpha^{(0)} = 5$ | $\omega \sim N(-3,5)$<br>$\omega_\alpha \sim N(-3,5)$<br>$\exp(\vartheta) = 0$<br>$\mu_v^{(0)} = 4$<br>$\mu_\alpha^{(0)} = 5$ |

**Supplementary Table 3. Bayesian model comparison in Experiment 1.** Exceedance probabilities across the four sessions for the three models described in Supplementary Table 2.

| | **HGF 1** | **HGF 2** | **HGF 3** |
|---|---|---|---|
| **Session 1** | 0.9863 | 0.0131 | 0.0006 |
| **Session 2** | 0.8608 | 0.0270 | 0.1123 |
| **Session 3** | 0.8166 | 0.1782 | 0.0053 |
| **Session 4** | 0.5117 | 0.4686 | 0.0197 |

**Supplementary Table 4.** The effect of model parameters in Experiment 1 on the participant-level random slope from the model predicting singed updates from signed prediction errors. Values of N = 45 participants and all four sessions were included. All variables were scaled.

| $B_U \sim \omega + \vartheta + \omega_\alpha$ | **Estimate** | **Std. Error** | **t value** | **Pr(>\|t\|)** |
|---|---|---|---|---|
| $(Intercept)$ | 0 | 0.038 | 0 | 0.999 |
| $\omega$ | 0.795 | 0.053 | 14.884 | <0.001 |
| $\vartheta$ | 0.54 | 0.059 | 9.108 | <0.001 |
| $\omega_\alpha$ | -0.008 | 0.065 | -0.121 | 0.904 |

**Supplementary Table 5.** The effect of model parameters in Experiment 1 on the participant-level random slope from the model predicting behavioral learning rates from variance block (factor). Values of N = 45 participants and all four sessions were included. All predictor variables were scaled.

| $B_L \sim \omega + \vartheta + \omega_\alpha$ | **Estimate** | **Std. Error** | **t value** | **Pr(>\|t\|)** |
|---|---|---|---|---|
| (Intercept) | -0.068 | 0.083 | -0.814 | 0.417 |
| $\omega$ | 0.146 | 0.107 | 1.371 | 0.172 |

| | | | | |
|---|---|---|---|---|
| $\vartheta$ | -0.22 | 0.105 | -2.097 | 0.037 |
| $\omega_\alpha$ | 0.438 | 0.115 | 3.822 | <0.001 |

**Supplementary Table 6.** The effect of model parameters in Experiment 1 on the participant-level random slope from the model predicting confidence from absolute prediction errors. Values of N = 45 participants and all four sessions were included. All variables were scaled.

| $B_C \sim \omega + \vartheta + \omega_\alpha$ | **Estimate** | **Std. Error** | **t value** | **Pr(>\|t\|)** |
|---|---|---|---|---|
| (Intercept) | 0 | 0.071 | 0 | 0.999 |
| $\omega$ | 0.216 | 0.1 | 2.155 | 0.032 |
| $\vartheta$ | 0.356 | 0.111 | 3.202 | 0.002 |
| $\omega_\alpha$ | -0.058 | 0.121 | -0.483 | 0.63 |

**Supplementary Table 7.** Effects of PDI on model parameters across sessions in Experiment 1. To check the robustness or our results across sessions, we ran three models each predicting one model parameter with PDI and a factor for session as a fixed and random effect. We repeated the same process for parameters estimated without pooling as well as parameters estimated by one hierarchical model.

$\vartheta \sim PDI * session + (session|ID)$

| Par. estimation | Hierarchical | | | | No pooling | | | |
|---|---|---|---|---|---|---|---|---|
| | **Est.** | **2.5%** | **97.5%** | **P** | **Est.** | **2.5%** | **97.5%** | **P** |
| $B_{PDI\_session1}$ | 0.486 | 0.01 | 0.964 | 0.024 | 0.39 | -0.062 | 0.851 | 0.05 |
| $B_{PDI\_session2}$ | 0.393 | -0.141 | 0.932 | 0.066 | 0.401 | -0.127 | 0.94 | 0.068 |
| $B_{PDI\_session3}$ | 0.358 | -0.157 | 0.901 | 0.086 | 0.181 | -0.294 | 0.64 | 0.234 |
| $B_{PDI\_session4}$ | 0.864 | 0.227 | 1.508 | 0.005 | 0.792 | 0.283 | 1.27 | 0.002 |

$\omega \sim PDI * session + (session|ID)$

| Par. estimation | Hierarchical | | | | No pooling | | | |
|---|---|---|---|---|---|---|---|---|
| | **Est.** | **2.5%** | **97.5%** | **P** | **Est.** | **2.5%** | **97.5%** | **P** |
| $B_{PDI\_session1}$ | -0.202 | -0.44 | 0.038 | 0.951 | -0.508 | -0.891 | -0.126 | 0.996 |
| $B_{PDI\_session2}$ | -0.215 | -0.456 | 0.031 | 0.96 | -0.389 | -0.798 | 0.006 | 0.974 |
| $B_{PDI\_session3}$ | -0.117 | -0.339 | 0.113 | 0.853 | -0.105 | -0.359 | 0.174 | 0.787 |
| $B_{PDI\_session4}$ | -0.128 | -0.403 | 0.149 | 0.819 | -0.101 | -0.491 | 0.278 | 0.7 |

$\omega_\alpha \sim PDI * session + (session|ID)$

| Par. estimation | Hierarchical | | | | No pooling | | | |
|---|---|---|---|---|---|---|---|---|
| | **Est.** | **2.5%** | **97.5%** | **P** | **Est.** | **2.5%** | **97.5%** | **P** |
| $B_{PDI\_session1}$ | 0.231 | -0.167 | 0.669 | 0.131 | 0.369 | -0.085 | 0.822 | 0.054 |
| $B_{PDI\_session2}$ | -0.026 | -0.578 | 0.552 | 0.535 | -0.27 | -0.931 | 0.403 | 0.786 |
| $B_{PDI\_session3}$ | 0.007 | -0.527 | 0.579 | 0.489 | -0.016 | -0.502 | 0.482 | 0.526 |
| $B_{PDI\_session4}$ | -0.006 | -0.61 | 0.625 | 0.508 | 0.039 | -0.548 | 0.642 | 0.439 |

**Supplementary Table 8.** In Experiment 2, effects of model parameters in the "Unusual Experiences" s ubscale of the Short Scale for Measuring Schizotypy. All continuous variables were scaled. Effect sizes reported in the main text obtained by dividing the effect of interest by residual standard deviation $\sigma$.

| $SCZ_{UE} \sim \omega + \vartheta + \omega_\alpha + Age + IQ + Gender$ | | | | |
|---|---|---|---|---|
| | **Estimate** | **Est.Error** | **Q2.5** | **Q97.5** |
| Intercept | 0.031 | 0.072 | -0.112 | 0.169 |
| $\vartheta$ | 0.09 | 0.047 | -0.001 | 0.181 |
| $\omega_\alpha$ | -0.012 | 0.048 | -0.106 | 0.083 |
| $\omega$ | -0.045 | 0.048 | -0.137 | 0.049 |
| $Age$ | -0.211 | 0.047 | -0.307 | -0.119 |
| $IQ$ | -0.065 | 0.047 | -0.159 | 0.029 |
| $Gender$ | -0.057 | 0.097 | -0.243 | 0.131 |

**Supplementary Table 9.** In Experiment 2, effects of model parameters in the "Introvertive Anhedonia " subscale of the Short Scale for Measuring Schizotypy. All continuous variables were scaled. Effect siz es reported in the main text obtained by dividing the effect of interest by residual standard deviation $\sigma$.

| $SCZ_{IA} \sim \omega + \vartheta + \omega_\alpha + Age + IQ + Gender$ | | | | |
|---|---|---|---|---|
| | **Estimate** | **Est.Error** | **Q2.5** | **Q97.5** |
| Intercept | 0.089 | 0.071 | -0.046 | 0.226 |
| $\vartheta$ | 0.016 | 0.047 | -0.077 | 0.112 |
| $\omega_\alpha$ | -0.08 | 0.049 | -0.176 | 0.017 |
| $\omega$ | -0.071 | 0.049 | -0.166 | 0.023 |
| $Age$ | *-0.176* | *0.048* | *-0.271* | *-0.082* |
| $IQ$ | *0.01* | *0.046* | *-0.078* | *0.1* |
| $Gender$ | *-0.16* | *0.095* | *-0.346* | *0.023* |

**Supplementary Table 10.** In Experiment 2, effects of two subscales of schizotypal traits: "Unusual Experiences" (UE) and "Introvertive Anhedonia" on confidence rating s on the next trial predicted by the confidence rating on the previous trial and prediction error on the previous trials and its interaction with the group level variables. Prediction error variable was scaled both within group and participant (id), group level variables were scaled within group. Confidence ratings were shrunk by 5% and logit transformed. Total variance was calculated by the sum of all variance component. Effect sizes reported in the main text are obtained by dividing the effect of interest by the squared root of the total variance.

| $Confidence^{(t+1)}_{LogOdds} \sim Confidence^{(t)}_{LogOdds} + PE^{(t)}(SZC_{UE} + SCZ_{IA}) + (PE^{(t)} \mid id)$ | | | | |
|---|---|---|---|---|
| Total Variance = 1.213 | **Estimate** | **Est.Error** | **Q2.5** | **Q97.5** |

| | | | | |
|---|---|---|---|---|
| $Intercept$ | 0.187 | 0.028 | 0.128 | 0.241 |
| $Confidence_{LogOdds}^{(t)}$ | 0.471 | 0.002 | 0.467 | 0.476 |
| $PE^{(t)}$ | -0.381 | 0.012 | -0.405 | -0.358 |
| $SZC_{UE}$ | 0.183 | 0.03 | 0.124 | 0.241 |
| $SCZ_{IA}$ | -0.06 | 0.03 | -0.117 | -0.001 |
| $PE^{(t)}SZC_{UE}$ | 0.042 | 0.012 | 0.018 | 0.066 |
| $PE^{(t)}SZC_{IA}$ | 0.012 | 0.012 | -0.011 | 0.036 |

**Supplementary Table 11.** In Experiment 2, effects of model parameters in the "compulsive behavior and intrusive thought" factor. All continuous variables were scaled. Effect sizes reported in the main t ext obtained by dividing the effect of interest by residual standard deviation $\sigma$.

| $CIT \sim \omega + \vartheta + \omega_{\alpha} + Age + IQ + Gender$ | | | | |
|---|---|---|---|---|
| | **Estimate** | **Est.Error** | **Q2.5** | **Q97.5** |
| Intercept | -0.004 | 0.068 | -0.137 | 0.131 |
| $\vartheta$ | 0.133 | 0.043 | 0.048 | 0.217 |
| $\omega_{\alpha}$ | 0.019 | 0.045 | -0.069 | 0.108 |
| $\omega$ | -0.06 | 0.044 | -0.149 | 0.027 |
| $Age$ | -0.318 | 0.045 | -0.407 | -0.231 |
| $IQ$ | -0.207 | 0.045 | -0.294 | -0.118 |
| $Gender$ | 0.007 | 0.093 | -0.179 | 0.194 |

**Supplementary Table 12.** In Experiment 2, effects of model parameters in the "anxious-depression" f actor. All continuous variables were scaled. Effect sizes reported in the main text obtained by dividing the effect of interest by residual standard deviation $\sigma$.

| $AD \sim \omega + \vartheta + \omega_{\alpha} + Age + IQ + Gender$ | | | | |
|---|---|---|---|---|
| | **Estimate** | **Est.Error** | **Q2.5** | **Q97.5** |
| Intercept | 0.155 | 0.07 | 0.018 | 0.293 |
| $\vartheta$ | -0.005 | 0.046 | -0.095 | 0.086 |
| $\omega_{\alpha}$ | -0.106 | 0.048 | -0.2 | -0.013 |
| $\omega$ | -0.122 | 0.048 | -0.216 | -0.029 |
| $Age$ | -0.221 | 0.047 | -0.313 | -0.128 |
| $IQ$ | -0.01 | 0.047 | -0.1 | 0.082 |
| $Gender$ | -0.281 | 0.094 | -0.465 | -0.1 |

**Supplementary Table 13.** In Experiment 2, effects of three factors of the factor analysis on confidence rating s on the next trial predicted by the confidence rating on the previous trial and prediction error on the previous trials and its interaction with the group level variables. Prediction error variable was scaled both within group and participant (id), group level variables were scaled within group. Confidence ratings were shrunk by 5% and logit transformed. Total variance was calculated by the sum of all variance component. Effect sizes reported in the main text are obtained by dividing the effect of interest by the squared root of the total variance.

| $Confidence_{LogOdds}^{(t+1)} \sim Confidence_{LogOdds}^{(t)} + PE^{(t)}(CIT + AD + SW) + (PE^{(t)} \mid id)$ | | | | |
|---|---|---|---|---|
| Total Variance = 1.201 | **Estimate** | **Est.Error** | **Q2.5** | **Q97.5** |
| $Intercept$ | 0.186 | 0.026 | 0.133 | 0.238 |
| $Confidence_{LogOdds}^{(t)}$ | 0.471 | 0.002 | 0.467 | 0.476 |
| $PE^{(t)}$ | -0.381 | 0.011 | -0.404 | -0.359 |
| $CIT$ | 0.264 | 0.031 | 0.202 | 0.327 |
| $AD$ | -0.118 | 0.034 | -0.185 | -0.054 |
| $SW$ | -0.035 | 0.036 | -0.107 | 0.035 |
| $PE^{(t)}CIT$ | 0.07 | 0.013 | 0.044 | 0.096 |
| $PE^{(t)}AD$ | 0.009 | 0.014 | -0.018 | 0.038 |
| $PE^{(t)}SW$ | -0.033 | 0.015 | -0.063 | -0.004 |

**Supplementary Table 14.** In Experiment 3, effects of model parameters for both avoid and seek sessi ons on positive symptoms measured by the Brief Psychiatric Rating Scale. All continuous variables we re scaled. Effect sizes reported in the main text obtained by dividing the effect of interest by $\sigma$.

| $BPRS \sim \eta_{avoid} + \omega_{avoid} + \vartheta_{avoid} + \omega_{\alpha,avoid} + \eta_{seek} + \omega_{seek} + \vartheta_{seek} + \omega_{\alpha,avoid} + Age + IQ + Gender$ | | | | |
|---|---|---|---|---|
| $\sigma = 1.20$ | **Estimate** | **Est.Error** | **Q2.5** | **Q97.5** |
| Intercept | 2.114 | 0.124 | 1.87 | 2.354 |
| $Age$ | 0.094 | 0.125 | -0.153 | 0.344 |
| $IQ$ | 0.086 | 0.126 | -0.166 | 0.334 |
| $Gender$ | -0.048 | 0.257 | -0.562 | 0.466 |
| $\eta_{avoid}$ (log noise) | 0.022 | 0.126 | -0.224 | 0.264 |
| $\omega_{avoid}$ | -0.035 | 0.133 | -0.29 | 0.232 |
| $\vartheta_{avoid}$ | -0.006 | 0.135 | -0.268 | 0.259 |
| $\omega_{\alpha,avoid}$ | 0.058 | 0.128 | -0.193 | 0.31 |
| $\eta_{seek}$(log noise) | 0.145 | 0.131 | -0.106 | 0.406 |
| $\omega_{seek}$ | -0.098 | 0.134 | -0.363 | 0.165 |
| $\boldsymbol{\vartheta_{seek}}$ | **0.224** | **0.127** | **-0.023** | **0.476** |
| $\omega_{\alpha,seek}$ | -0.2 | 0.137 | -0.467 | 0.068 |

**Supplementary Table 15.** In Experiment 3, effects of model parameters for both avoid and seek sessions on negative symptoms measured by the Scale for the Assessment of Negative Symptoms. All continuous variables were scaled. Effect sizes reported in the main text obtained by dividing the effect of interest by $\sigma$.

$$BPRS \sim \eta_{avoid} + \omega_{avoid} + \vartheta_{avoid} + \omega_{\alpha,avoid} + \eta_{seek} + \omega_{seek} + \vartheta_{seek} + \omega_{\alpha,avoid} + Age + IQ + Gender$$

| $\sigma = 0.62$ | **Estimate** | **Est.Error** | **Q2.5** | **Q97.5** |
|---|---|---|---|---|
| Intercept | 1.425 | 0.065 | 1.297 | 1.553 |
| $Age$ | 0.225 | 0.066 | 0.097 | 0.353 |
| $IQ$ | 0.063 | 0.063 | -0.062 | 0.188 |
| $Gender$ | -0.136 | 0.139 | -0.408 | 0.138 |
| $\eta_{avoid}$ (log noise) | -0.009 | 0.065 | -0.137 | 0.118 |
| **$\omega_{avoid}$** | **-0.137** | **0.07** | **-0.274** | **0.002** |
| $\vartheta_{avoid}$ | -0.007 | 0.071 | -0.148 | 0.133 |
| **$\omega_{\alpha,avoid}$** | **-0.184** | **0.066** | **-0.316** | **-0.055** |
| **$\eta_{seek}$(log noise)** | **-0.14** | **0.068** | **-0.273** | **-0.002** |
| $\omega_{seek}$ | 0.006 | 0.068 | -0.127 | 0.143 |
| $\vartheta_{seek}$ | 0.052 | 0.066 | -0.079 | 0.184 |
| $\omega_{\alpha,seek}$ | 0.015 | 0.069 | -0.118 | 0.149 |

#### Supplementary Figures

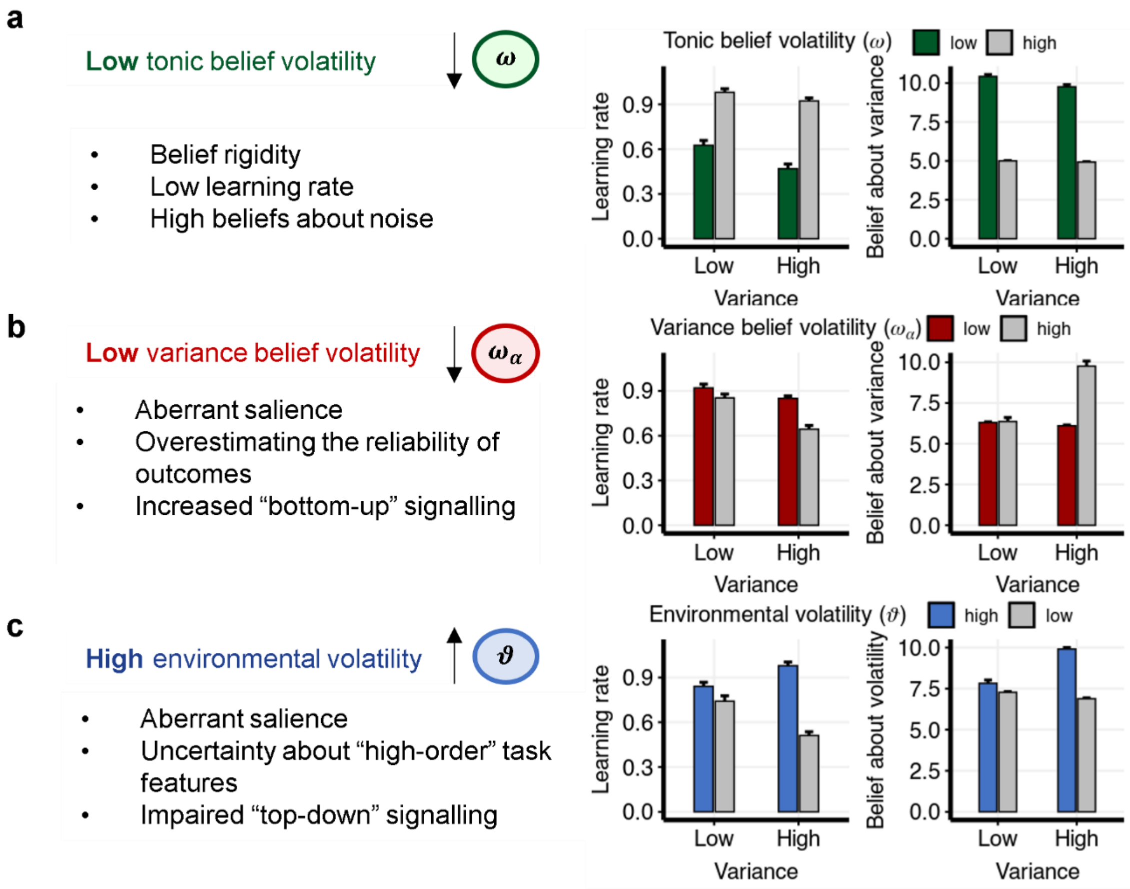

**Supplementary Figure 1. Model simulations.** Model simulations of two agents with different parameter sets. **a**, Different $\omega$ parameters and fixed and low values for $\omega_\alpha$ and $\vartheta$. **b**, Different $\omega_\alpha$ and fixed $\omega$ and $\vartheta$. **c**, Different $\vartheta$ and fixed $\omega$ and $\omega_\alpha$. Learning rate refers to the ratio of action update and signed prediction error. Bar plots are means across blocks for each agent with error bars representing standard errors. Simulations are done with 480 trials for two agents (one agent per parameter set). Through this, we can define several computational patterns of belief updating. When comparing simulated behavior and belief trajectories of an agent with lower (compared to higher) tonic belief volatility the model implies lower prior precision and therefore lower precision-weighted learning rate ($\psi_1$). Therefore, the behavioral learning rate is reduced across blocks with both low and high outcome variance. Furthermore, large deviations from expected outcomes are explained as noise, leading to high beliefs about outcome variance. In contrast, in the case of a lower outcome variance volatility simulations show an increased learning rate, particularly in blocks with high variance, resulting from a failure to update beliefs about the noise in high variance blocks. When environmental volatility is higher , higher outcome variance is interpreted as increased volatility, leading again to higher learning rates in the high variance block. Both latter computational patterns can be perceived as related to aberrant salience attribution; however, they might be implemented in distinct neural circuits.

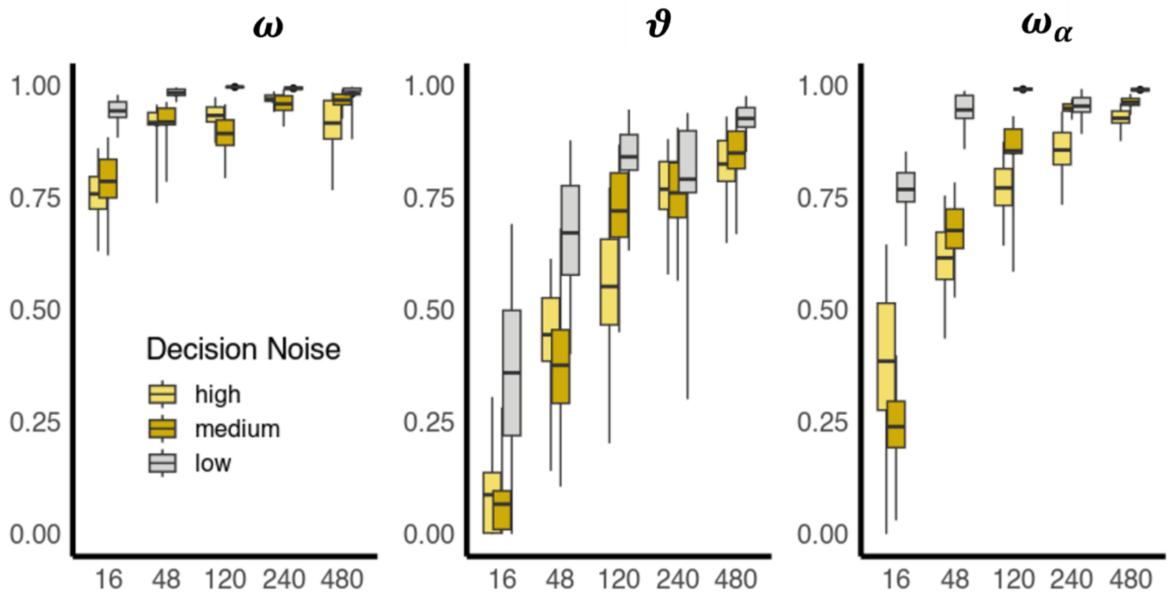


**Supplementary Figure 2. Parameter recovery.** We simulated behavior for n = 50, with parameters drawn from the prior distribution and re-estimated the parameters for various trial lengths and three decision noise levels.

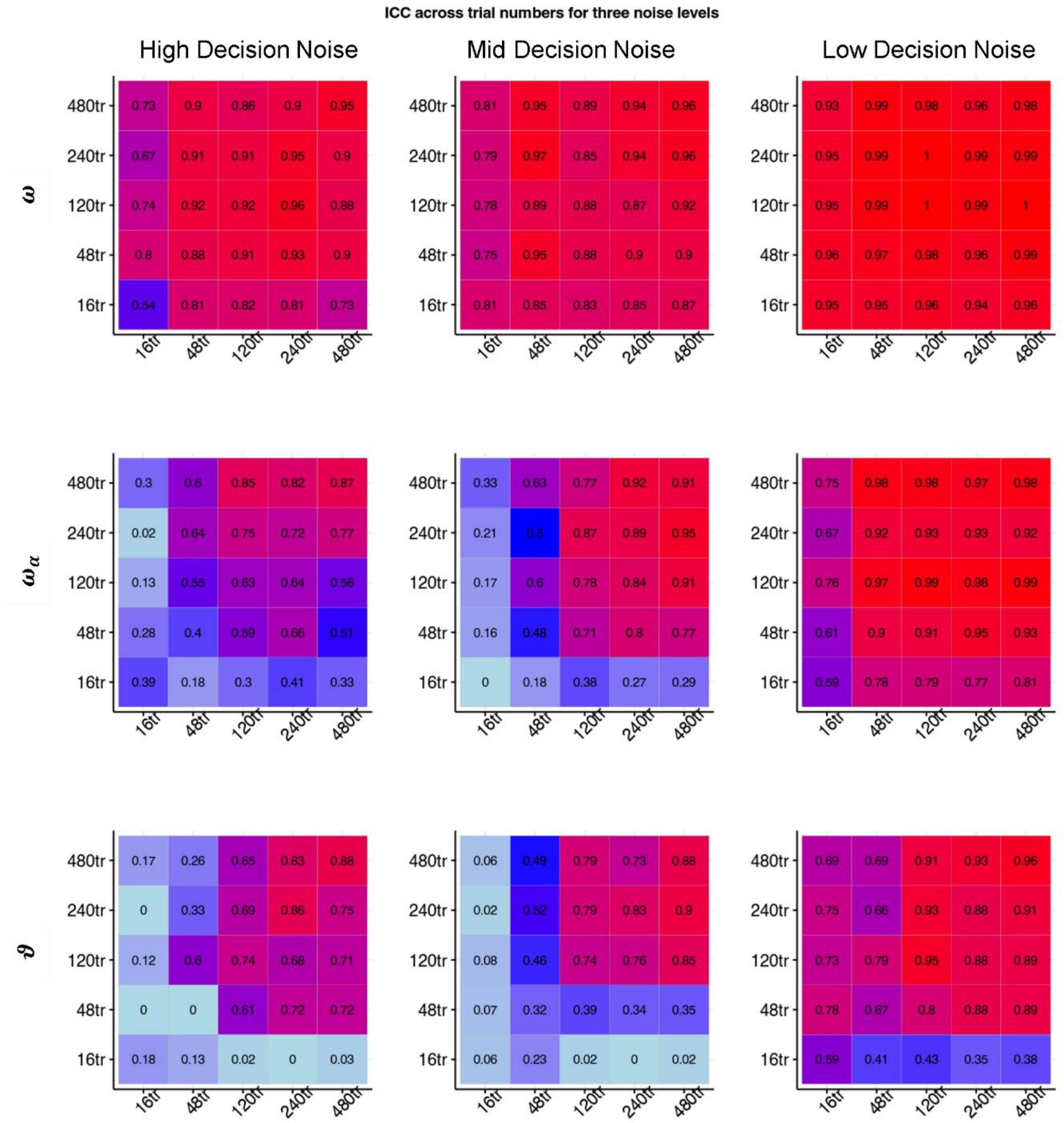


**Supplementary Figure 3.** Simulating ICCs across trial numbers for three different levels of decision noise. tr – Trials

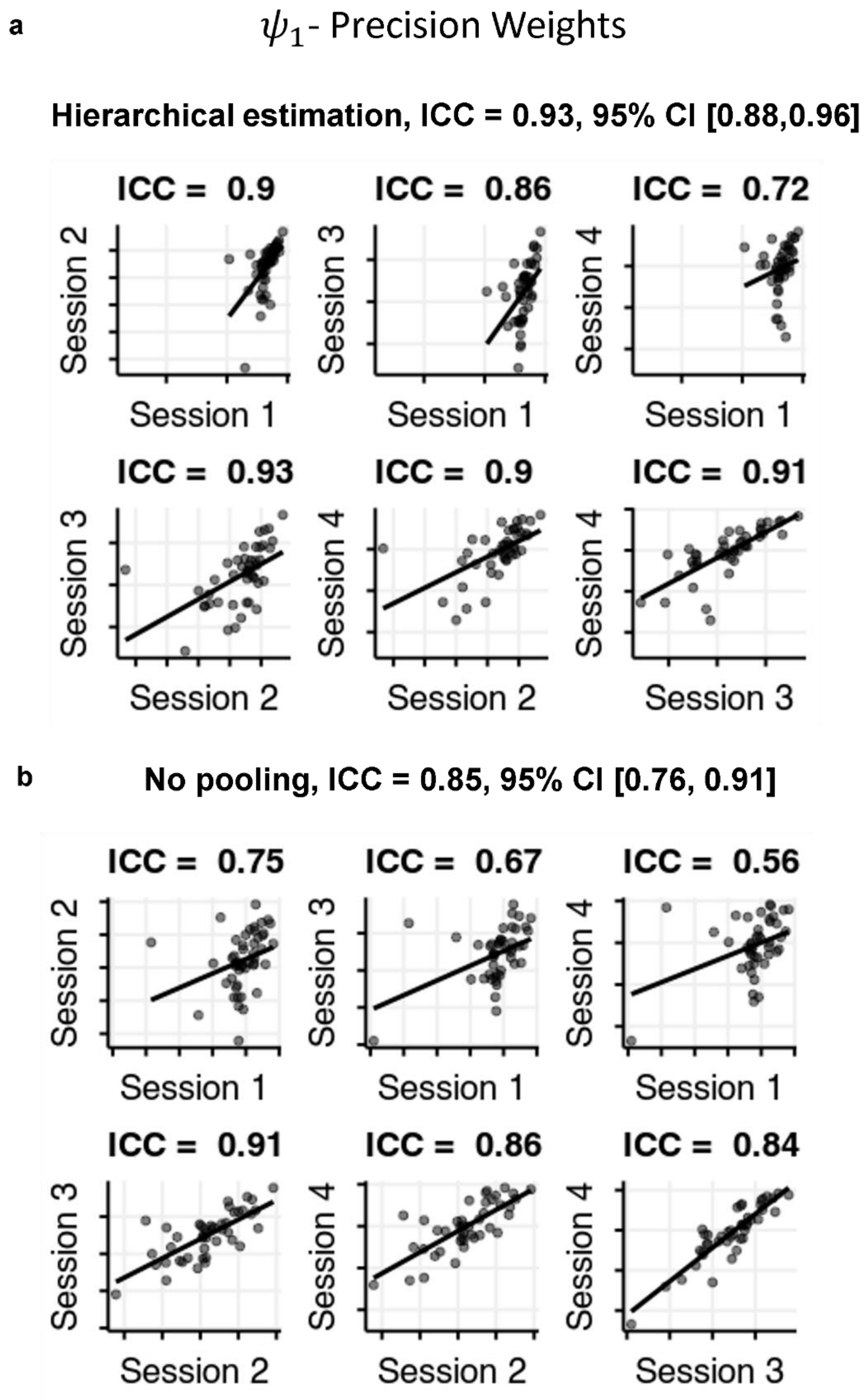


**Supplementary Figure 4.** ICCs and regression lines for average precision weights for parameter estimation with (a) and without (b) pooling.

**a** $\mu_\alpha$- Beliefs about outcome variance

**Hierarchical estimation, ICC = 0.92, 95% CI [0.88,0.95]**

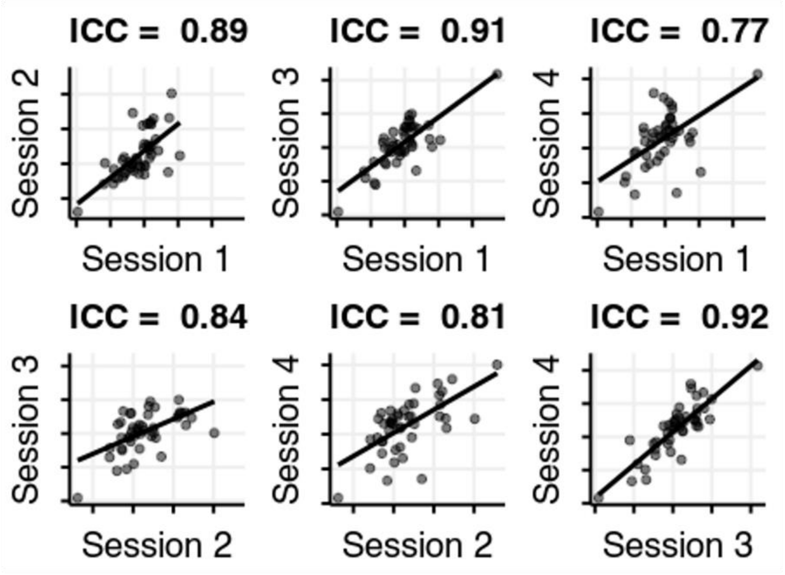


**b**

**No pooling, ICC = 0.86, 95% CI [0.77, 0.91]**

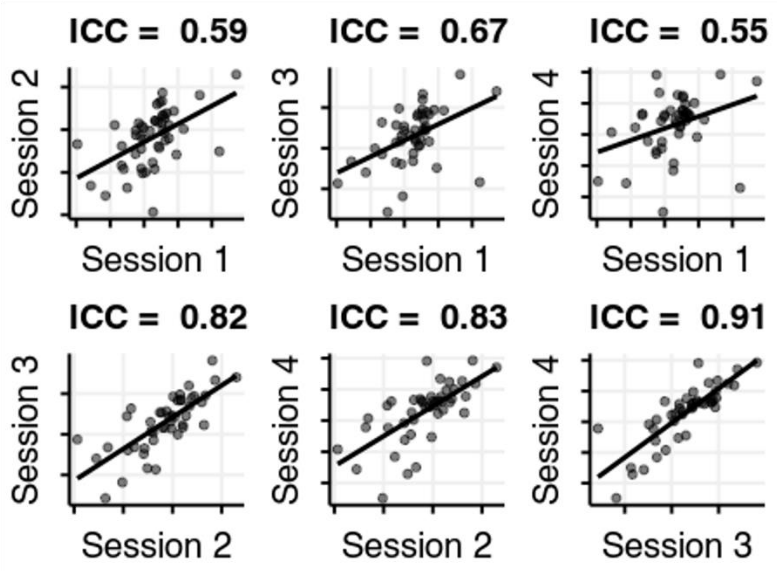


**Supplementary Figure 5.** ICCs and regression lines for average beliefs about variance for parameter estimation with (a) and without (b) pooling.

**a** $\mu_v$ - Beliefs about environmental volatility

**Hierarchical estimation, ICC = 0.92, 95% CI [0.87,0.95]**

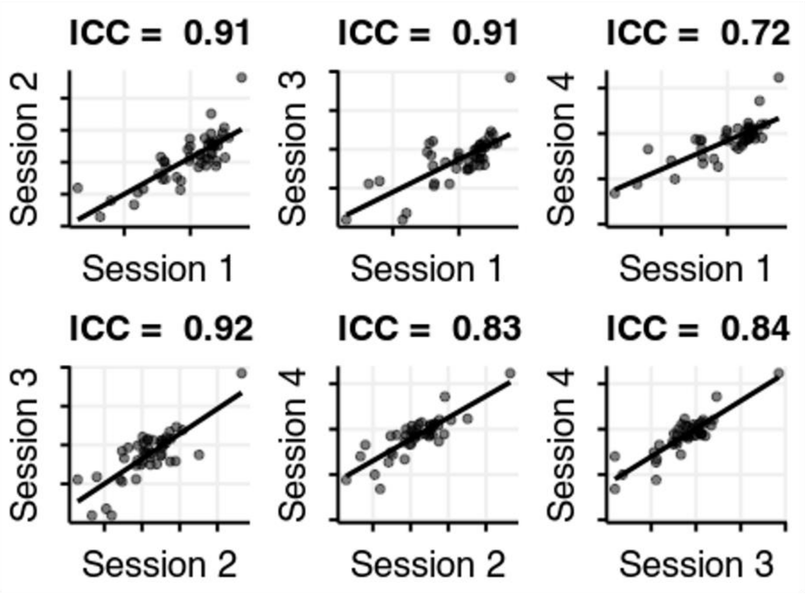


**b**

**No pooling, ICC = 0.47, 95% CI [0.16, 0.68]**

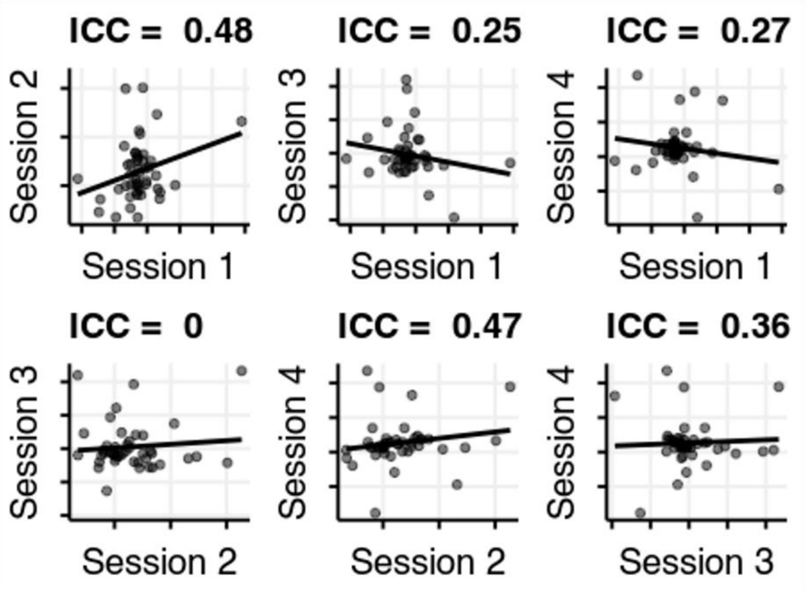


**Supplementary Figure 6.** ICCs and regression lines for average beliefs about volatility for parameter estimation with (a) and without (b) pooling across sessions.

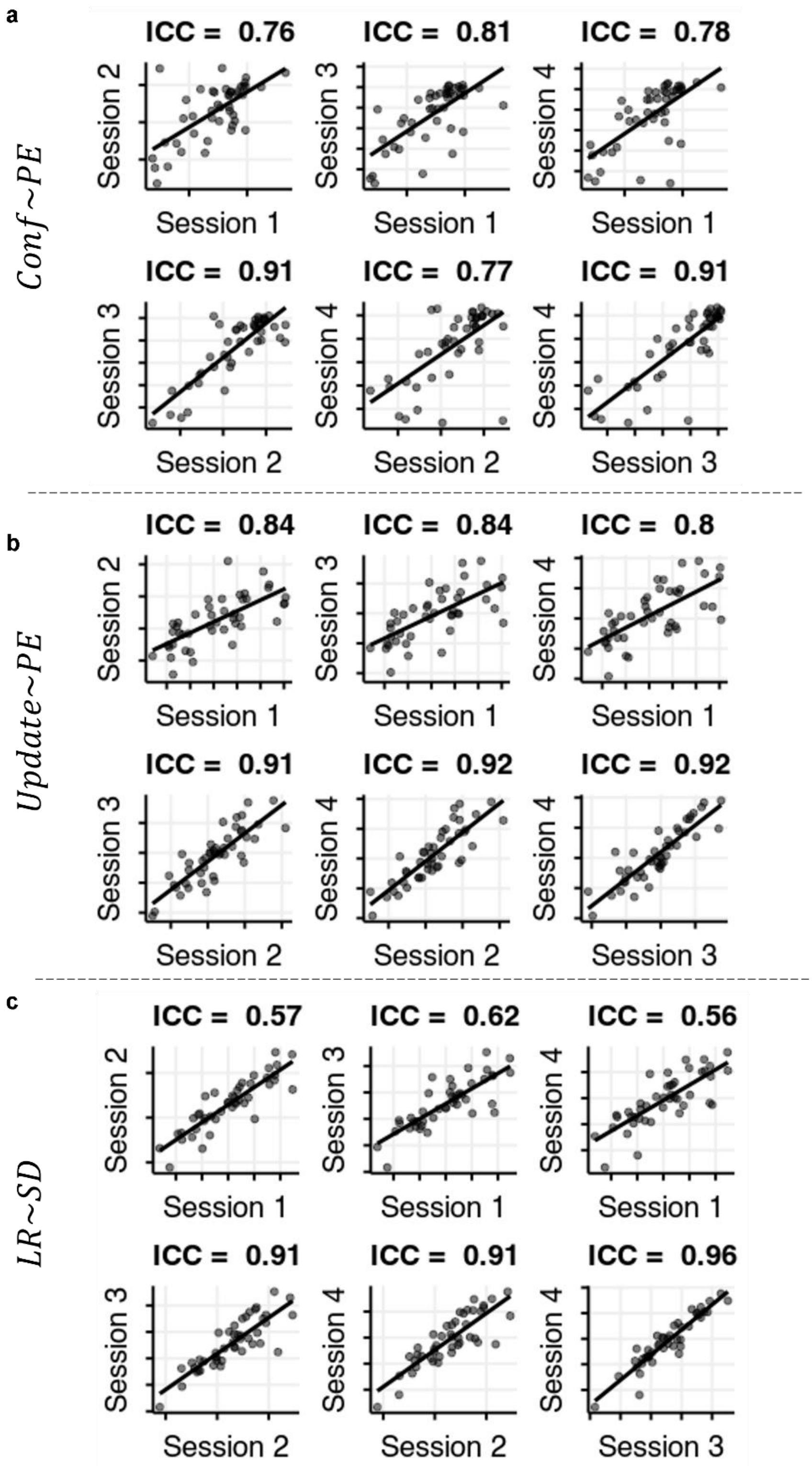


**Supplementary Figure 7.** ICCs and regression lines for behavioral patterns (a) $B_c$, (b) $B_U$, and (c) $B_L$ defined by the random slopes of three models, estimated per session.

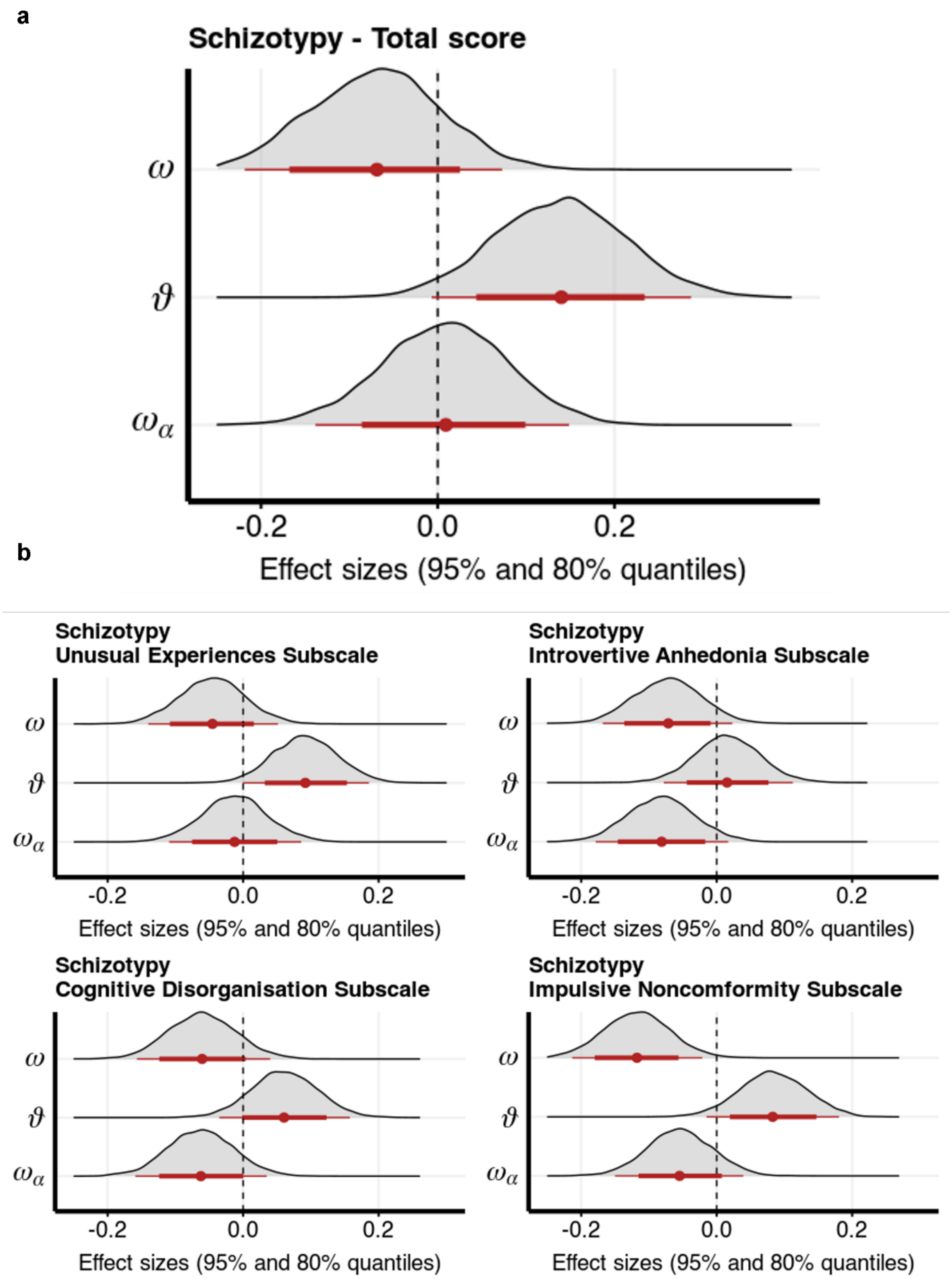


**Supplementary Figure 8. Modelling results Seow and Gillan (2020). a** Total score of the Short Scale for measuring Schizotypy predicted by the three parameters controlling for age, gender and IQ. **b**, All subscales of the Short Scale for measuring Schizotypy were predicted by the three parameters controlling for age, gender and IQ. Plotting posterior distributions of effects of each model parameter, over means and CrIs of effect sizes.

### Supplementary Methods

#### Behavioural Analysis

Variables that were only positive were translated by 1 (to avoid dividing by zero), and log-transformed (e.g. learning rate, absolute (unsigned) update, and PDI). Where possible, all within subject variables were used as random effects. In all linear models, all continuous variables were scaled. For Bayesian models we report Credibility intervals (CrI) and the probability of the posterior interval that lies above (or below) zero. Effect sizes for linear models with random effects were calculated by dividing the regression coefficient of interest with the square of the total variance, calculated as a sum of residual variance and the variance of all random effects (Nalborczyk, Batailler, Loevenbruck, Vilain, & Bürkner, 2019).

#### Hierarchical Bayesian Model definition

Recent work has shown that hierarchical inference can improve reliability scores (Waltmann, Schlagenhauf, & Deserno, 2022), because it reduces overfitting (McElreath, 2018), pools information across different levels (testing sessions and participants) and allows for estimation of both participant parameters as well as group-level relevant statistics (such as the ICC scores) in one inferential step.

The participant level parameters for session 1 ($\omega$, $\omega_\alpha$, $\vartheta$, and $\eta$), were modelled as correlated (multivariate) effects with

$$\begin{pmatrix} \omega_{\mathrm{id}} \\ \omega_{\alpha,\mathrm{id}} \\ \vartheta_{\mathrm{id}} \\ \eta_{\mathrm{id}} \end{pmatrix} \sim \mathrm{MVNormal}(\boldsymbol{\mu}, \boldsymbol{S}), \qquad (1)$$

$$S = \boldsymbol{\sigma^b R \sigma^b},$$

where $\boldsymbol{S}$ is the covariance matrix and $\boldsymbol{\mu}$ is the vector of mean values for each parameter. The matrix $\boldsymbol{S}$ was factored into a diagonal matrix with between subject standard deviations of the parameters ($\sigma^b$) and the correlation matrix $\boldsymbol{R}$ (Bürkner, 2017; McElreath, 2018). For each parameter (e.g. $\omega$) for participant $id$ and session $s$ we therefore defined

$$\omega_{id,s} = \mu_\omega + b_s + r_{id} + r_{id,s} ,$$

where $\mu_\omega$ is the overall mean, $b_s$ is the overall session (fixed) effect, $r_{id}$ is the participant level intercept, and $r_{id,s}$ is the participant level effect of session. With the following prior structure:

$$r_{id,s} \sim N(0, \sigma_\omega^w),$$

$$b_s \sim N(0,1)$$

$$\sigma_\omega^w \sim HalfCauchy(0,2),$$

$$\sigma_\omega^b \sim HalfCauchy(0,2),$$

$$R \sim LKJcorr(2),$$

$$\mu_\omega, \mu_\eta \sim N(0,1),$$

$$\mu_{\omega_\alpha}, \mu_\vartheta \sim N(-6,1)$$

The intraclass correlation (*ICC*) for the parameters was defined as the ratio of between-subjects variance ($\sigma_b^2$) versus total variance:

$$ICC = \frac{\left(\sigma^b\right)^2}{(\sigma^b)^2 + (\sigma^w)^2}.$$

#### Predictive Inference task

##### *Experiment 1*

After a change point the new mean was drawn from a uniform distribution on the [10,90] interval and the standard deviation could be either high (15) or low (5). Participants are required to express their confidence in their choice by holding the response button for longer (for a max of 1.4 seconds). Participants were reminded several times throughout the task to pay attention to the confidence ratings, with a text message flashed on the screen during the prediction phase of the next trial.

The task was implemented in Psychtoolbox in Matlab (Mathworks). We embedded the task within a story and incentivized accuracy. Participants were told to predict the amount of gold (indicated by a yellow bar) an alien will bring back from a mine on each trial with their arrow keys. Whereas the alien would usually bring back similar amounts of gold from one mine, it might also change mines once and a while. Participants were trained in a sample of mines that differed both in the mean amount of gold they produced as well as the variance. The participants were told that the amount of money they earned on each trial depended on the size of the yellow bar and how close they were to the actual outcome. A full bar was indicative of € 1 of potential gain. Participants first underwent a 20-minute training session where they would experience low and high variance blocks as well as several mean shifts, where the changes of mines were made obvious through visual cues.

In each session participants played 240 trials of the task. Four different trajectories were used for the four sessions and the trajectories were the same for all participants. The trajectories were selected randomly but with a fixed number of reversals. Furthermore, we conditioned the selection of the trajectories on the model. To maximize test-retest reliability, we selected trajectories where the parameters of an ideal Bayesian observer (given the priors in Table 2) were not too far away from each other (i.e. the Mahalanobis distance was below the 5$^{th}$ percentile).

##### *Experiment 2:*

In contrast to the task used in Experiment 1, the variance of responses did not change, but the volatility did. In high volatility blocks of trials, the probability for a change on each trial was 0.125, and in low volatility blocks it was 0.025. In this version of the task participants were told to catch a particle flying from an invisible canon from the center to the edge of a large circle. Participants would gain points when they caught the particle and lose points otherwise. All participants played 300 trials of the task, with one volatility condition across 150 trials. In contrast to Experiment 1, each participant’s trajectory was randomly generated. The standard deviation around the mean bucket position was 12 from a 360-point scale (which compares to a standard deviation of 3.33 on a scale from 1 to 100 used in Experiment 1).

##### *Experiment 3*

The version of the task was comparable to that in Experiment 2 and to other studies published previously (Nassar, Wilson, Heasly, & Gold, 2010). Participants were told, they need to catch bags that are dropped from a helicopter, whereby they cannot see the helicopter and need to infer its position based on the dropped bags (McGuire, Nassar, Gold, & Kable, 2014). The probability of the helicopter changing its position on each trial was 0.125 (as in the high volatility block of Experiment 2 and comparable to the volatility in Experiment 1), and the standard deviation was 20 on a 300-point scale (which compares to a standard deviation of 6.67 on a scale from 1 to 100 used in Experiment 1). Contrary to both Experiment 1 and 2, participants' performance was incentivized in two different ways across two different task sessions, each lasting for 100 trials. In the appetitive condition, participants could earn points and in the aversive condition, participants would have points taken away from their initial endowment in a performance contingent way.

##### Two-step Task

The task was based on the version from Kool, Cushman, & Gershman (2016), and was slightly adapted (Mikus et al., 2022). Participants are required to earn points in a dynamically changing environment whereby each trial consists of two steps or stages. In the first stage, participants see one of two possible starting scenarios, each featuring a pair of spaceships that fly deterministically to one of two second stages that feature planets, where they encounter an alien that gives them points ranging from -4 to +5. How many points each alien gives changes throughout the experiment according to a Gaussian random walk. The deterministic transition between the spaceships and the planets stays the same throughout the experiment and participants are explicitly instructed about it. This represents the participants' "knowledge about the regularities in the world". In this version of the task, referring to this mapping when making decisions leads to higher payoff in the task (Kool et al., 2016).
